# Locus 9p21.3 CAD Risk Across Diverse Populations: LD and Allelic Heterogeneity Explain Variation Across Ancestries and Reduced Replication in Africans

**DOI:** 10.64898/2026.01.02.25343215

**Authors:** Hasan Alkhairo, Satoshi Koyama, Kruthika Iyer, Austin T Hilliard, Pik Fang Kho, Shoa Clarke, Firdous M Abdulwahab, Themistocles L Assimes, Julie A Lynch, Michèle Ramsay, Human Heredity and Health in Africa (H3Africa), Africa-Wits INDEPTH (AWI-Gen), VA Million Veteran Program (MVP), Kyong-Mi Chang, Philip S Tsao, Fowzan S Alkuraya, Kaoru Ito, Neil Risch, Catherine Tcheandjieu

**Affiliations:** Gladstone Institutes of Data Science and Biotechnology, Gladstone Institute, 1650 Owens street, San Francisco, CA, 94158, USA; VA Palo Alto Health Care System, Palo Alto, CA, USA; Program in Medical and Population Genetics and the Cardiovascular Disease Initiative, Broad Institute of Harvard and MIT, Cambridge, MA; Department of Medicine, Stanford University, CA, USA; King Faisal Specialist Hospital & Research Centre, Riyadh, Saudi Arabia; School of Medicine, Epidemiology, University of Utah, UT, USA; Salt Lake City VA Medical Center, Salt Lake City, UT, USA; Sydney Brenner Institute for Molecular Bioscience, University of the Witwatersrand, Johannesburg, South Africa; A list of all MVP contributions can be found in the Data Supplement; Corporal Michael J. Crescenz VA Medical Center, Philadelphia, PA, USA; Department of Medicine, University of Pennsylvania, Philadelphia, PA, USA; Laboratory for Cardiovascular Genomics and Informatics, RIKEN Center for Integrative Medical Sciences, 1-7-22 Suehiro-cho, Tsurumi-ku, Yokohama, 230-0045, Japan; Department of Advanced Biomedical Data Sciences, Chiba University Graduate School of Medicine, 1-8-1 Inohana, Chuo-ku, Chiba, 260-8670, Japan; Department of Epidemiology and Biostatistics, University of California San Francisco, CA, USA; Institute for Human Genetics, University of California San Francisco, CA 94143

## Abstract

**Background:** The 9p21.3 locus was the first genome-wide significant signal for coronary artery disease (CAD) and replicates across multiple non-African populations, yet is absent in African ancestry cohorts. We hypothesized that ancestry-specific linkage disequilibrium (LD) and haplotype structure, rather than allele frequency or power alone, explain this discrepancy.

**Methods:** We analyzed multi-ancestry data from European, East Asian, South Asian, Middle Eastern, African, and Admixed American groups. Within each ancestry, we performed CAD common-variant associations at 9p21.3, rare-variant tests in whole-genome-sequenced Europeans, local-ancestry inference (LAI) stratified associations, conditional and haplotype analyses, and pleiotropy assessments.

**Results:** CAD associations at 9p21.3 were robust in Europeans, East Asians, Middle Eastern, and Admixed Americans ancestry groups, nominal in South Asians, and absent in Africans despite similar allele frequencies for lead-associated SNPs in other ancestry groups. LAI-stratified analyses showed strong signals in African heterozygotes carrying 9p21.3 European chromosomes but not in 9p21.3-homozygous Africans. The association is due to highly common variants; rare variants did not explain the locus signal. LD blocks were extended in non-Africans but fragmented into smaller blocks in Africans, with marked divergence by fixation index. Fine-mapping identified multiple independent signals, including an East Asian-specific haplotype absent in Europeans and Africans. Hamming-distance analyses revealed that risk alleles are dispersed across multiple haplotypes in homozygous African groups, consistent with weaker LD and restricting risk variation at this locus in that group. PheWAS mirrored these ancestry-specific patterns across metabolic traits.

**Conclusions:** The non-replication of 9p21.3 in African ancestry is not due to absent risk alleles or poorer imputation of missing risk alleles due to weaker LD; rather, it reflects greater haplotype diversity that disperses risk alleles across configurations, attenuating case-control contrast. These findings illustrate how ancestry-specific LD and multi-causal haplotype architecture modulate association detectability, with broader implications for initial association detection and fine-mapping.

## Introduction

Locus 9p21.3 is the first genome wide locus identified for coronary artery disease (CAD) by genome-wide association studies (1,2). The locus was first identified in Europeans in a study of 2000 cases and 2000 controls (1) and since then, has been replicated in East Asian, South Asian, Middle Eastern, and Admixed American cohorts (1–4). Additionally, 9p21.3 appears to be highly pleiotropic, showing association with various diseases including diabetes, metabolic syndrome, various cancers, and more recently lipid levels (5). Since its discovery numerous studies have sought to elucidate its functional role in CAD pathogenesis (5–8). Within the 9p21.3 locus focal genes included *CDKN2A*, *CDKN2B*, and a .45 Mb long noncoding RNA (lncRNA) *ANRIL* (*CDKN2B-AS1*), which harbors the strongest association signal across the 0.45 Mb region.

Despite its reproducible association in European, East Asian, Middle Eastern, Admixed American, and South Asian populations, the 9p21.3 signal is notably absent in African ancestry groups—even in well-powered studies where sample size is unlikely to be a limiting factor (4,2,3).

While functional studies have grown increasingly sophisticated and have brought us closer to understanding the role of 9p21.3 in CAD, the persistent lack of replication of the 9p21.3 signal in African ancestry populations and its explanation has become an important knowledge gap. This inconsistency highlights the need to consider how population-level features, such as linkage disequilibrium (LD) and minor allele frequency (MAF), interact with local genetic architecture to influence both the detection and interpretation of association signals. The need to examine ancestry-specific genetic architecture in complex diseases has been particularly emphasized in the context of polygenic risk score (PRS) transferability, where differences in LD and MAF across populations significantly affect PRS performance across groups (9). While population-level factors are often discussed in the context of risk prediction, their role in modulating association signals, and in shaping causal architecture at loci such as 9p21.3, remains insufficiently understood.

In this study, we conduct a comprehensive analysis of the 9p21.3 locus using data from multiple large-scale cohorts representing diverse ancestries. We examine association with SNPs at 9p21.3 within each ancestry group independently, perform fine-mapping analyses to resolve potential candidate causal variants, and investigate local linkage disequilibrium and haplotype structure using both targeted and expanded SNP sets. By integrating these approaches, we aim to clarify how the genetic architecture at 9p21.3 and its relationship to CAD risk vary by ancestry, and most importantly why the 9p21.3 signal is consistently absent in African ancestry groups and what this reveals about the interplay between genetic architecture and population structure.

## Results

### Association with CAD at the locus 9p21.3 across multiple populations

To investigate the genetic architecture of CAD at 9p21.3 across ancestries, we analyzed CAD association across multiple populations using data from various biobanks and cohorts, encompassing European (EUR), East Asian (EAS), South Asian (SAS), Middle Eastern (MID), African (AFR), and Admixed American (AMR) populations (detailed in Methods). Details on association testing, genetically inferred ancestry (GIA) assignments, and CAD sample sizes are outlined in Methods and Table S0. Variants crossed the genome-wide significance threshold (p < 5E-08) in EUR, EAS, AMR, and MID (Saudi) populations; surpassed the suggestive significance threshold (p < 1E-05) in SAS, and did not attain significance in African ancestry (Fig. 1, Tables S5-7,24-25). In all associations, the 9p21.3 signal peaks at a median position of 22.1 MB across all ancestries (Fig. 1), and a second cluster of significant variants is observed just upstream, between 21.69 Mb and 21.84 Mb with a maximum around 21.82 MB in EUR (Figure 1A). The peak signal at 22.1 MB had a risk allele frequency around 0.49 while; the signal at 21.82 Mb had a risk allele frequency around 0.46.

**Figure 1:**
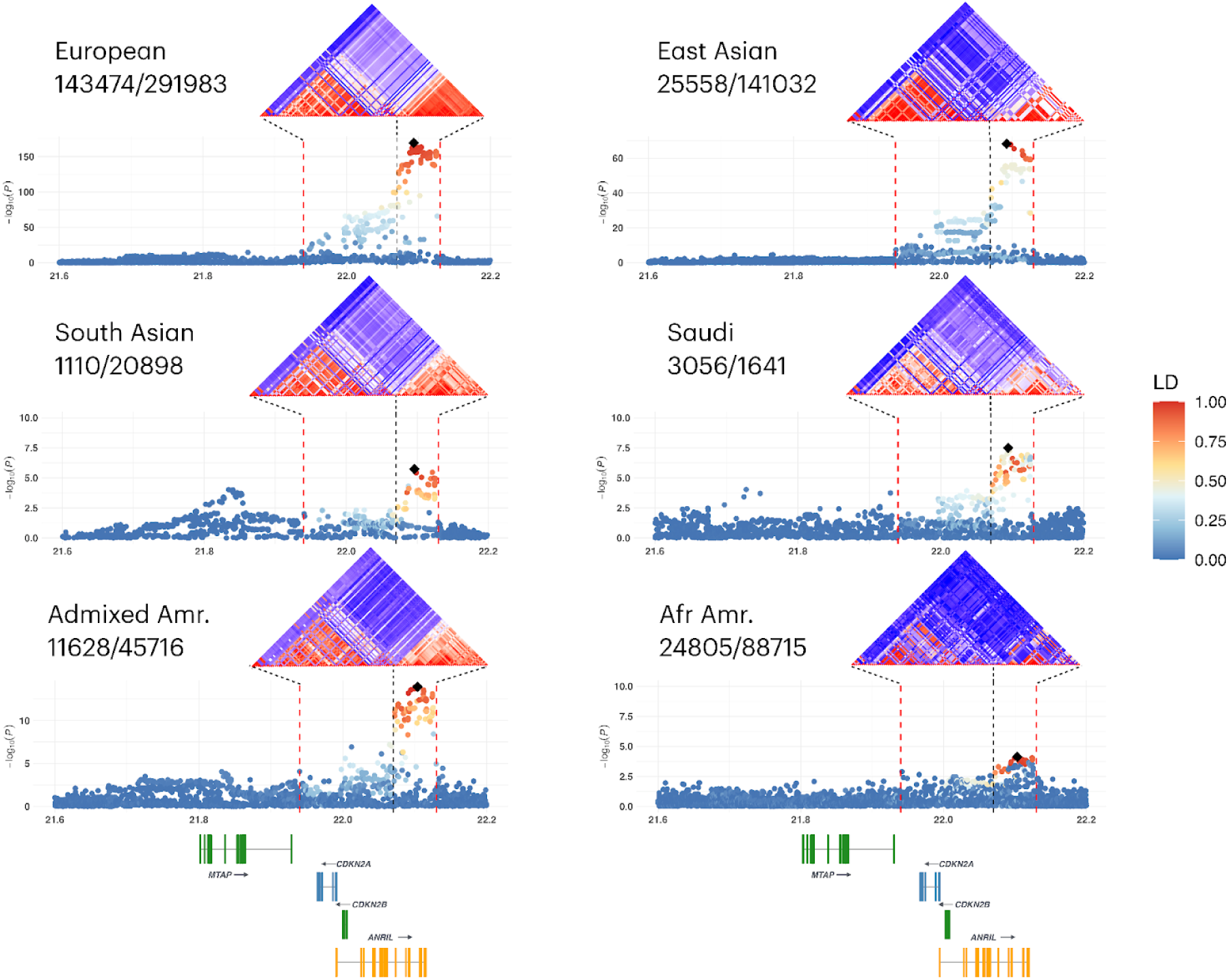
Locus zoom plots of 9p21.3 stratified by ancestry. Locus zoom plots are shown for each ancestry group, with case/control sample sizes indicated next to group labels. Linkage disequilibrium (LD) reflects in-sample r2 for each group, and all analyses followed a uniform preprocessing pipeline except for the South Asian analysis, which used publicly available summary statistics from Genes & Health. Black dashed vertical lines denote the primary association hotspot at 9p21.3, while the red vertical line marks the boundary between two distinct LD blocks. SNPs shown in the LD heatmaps reached genome-wide significance in at least one group and had a minor allele frequency >1% in all groups. Results for globally inferred ancestry groups: MVP European (European), BioBank Japan (East Asian), the King Faisal Specialty Hospital and Research Centre CAD study (Saudi), the globally inferred AMR group in MVP (Admixed American), and the globally inferred AFR group in MVP (African American). The South Asian association is based on summary statistics, and its corresponding LD was estimated using the genetically inferred South Asian cohort in UK Biobank.

To investigate the contribution of rare variants at the 9p21.3 locus we leveraged UK Biobank (UKBB) whole-genome sequencing data. Using REGENIE(10), we performed an association analysis incorporating rare variants in a group of 457,954 EUR individuals (Table S4). No rare SNPs reached genome-wide significance (Fig S1A), and lower-frequency variants (MAF < 2%) tend to have odds ratios that are either smaller or comparable to those of the most common variants (MAF ∼50%).

To refine ancestry-specific associations at 9p21.3, particularly in AFR and AMR individuals, we performed multi-local ancestry inference in the Million Veteran Program (MVP) using Gnomix (11) using EUR, AFR, NAT, EAS, SAS from 1000genome and HGDP panel as reference (see Methods). We reclassified MVP participants based on their ancestry inferred at the locus 9p21.3 intervals as heterozygous or homozygous for each of the reference populations as described in Methods (Fig. S8) and conducted association testing within these local ancestry-inferred (LAI) groups (Tables S0, Tables S18-S23).

Figure 2 shows the locuszoom plot of association in homozygous EUR (EUR|EUR), homozygous AFR (AFR|AFR), and homozygous NAT (NAT|NAT) representing individuals who were 9p21.3-wide homozygous EUR, AFR, and NAT, respectively. Furthermore, association in heterozygous groups for AFR (AFR|non-AFR) and NAT (NAT|non-NAT) are included, and finally a group with homozygous EUR chromosomes among GIA AFR (GIA-AFR:EUR|EUR) (individuals with global AFR ancestry inferred genome-wide). As expected, significant associations were observed in the homozygous European (EUR|EUR) group with the lead SNP (9:22093299:A:T) matching that observed in the EUR in MVP (OR=1.14, Table S3). In the heterozygous African (AFR|non-AFR, n-∼46,000), 11 SNPs reached genome-wide significance (Table S19).

**Figure 2:**
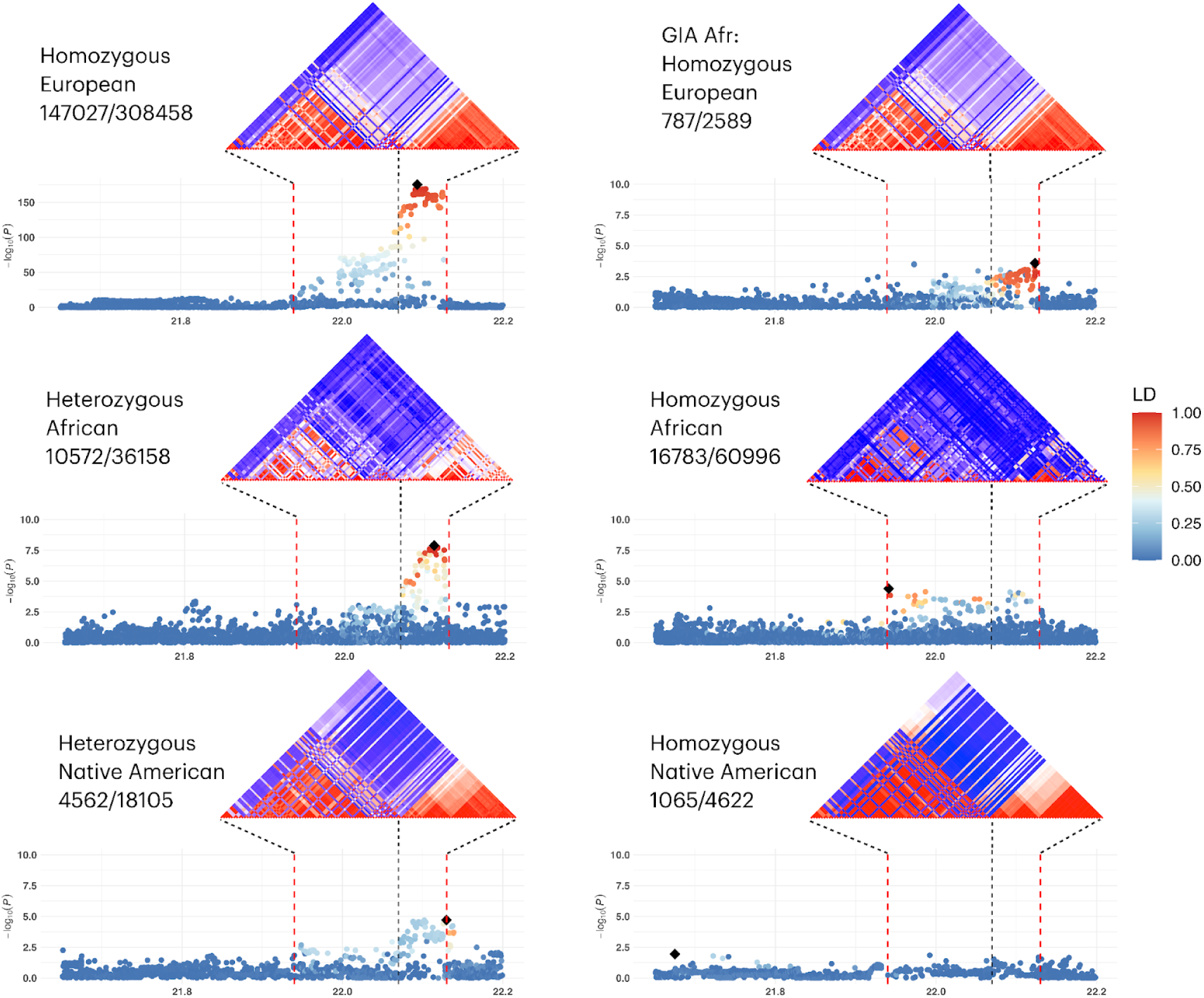
Locus zoom plots of 9p21.3 stratified by local ancestry in MVP. Locus zoom plots for locally inferred ancestry groups within MVP, with case/control sample sizes indicated next to group labels. Linkage disequilibrium (LD) reflects in-sample r^2^ for each group, and all analyses. Further details are described in Figure 1 legend. “GIA Afr: Homozygous Euro” refers to individuals globally inferred as African ancestry who are homozygous for European ancestry at 9p21.3.

The lead variant, an indel (9:22111584:T:TATTTG), exhibited an odds ratio of 0.88 and a p-value of 1.28 × 10⁻⁸, with a minor allele frequency (MAF) of 26% in the heterozygous group. Notably, the same variant had a MAF of 4% in the homozygous African group with no significant association (OR=.99, p=.77) and 48% in the homozygous European group (OR=.88, p=3.83E-160). All 11 genome-wide significant variants in the AFR heterozygous group appear to be co-inherited on EUR chromosomes, as indicated by their consistent minor allele frequency patterns: approximately 25% in heterozygous individuals, ∼3% in homozygous Africans, and ∼48% in homozygous Europeans. In the homozygous African group (AFR|AFR, n = ∼77K), 4 variants show nominal significance (Table S18) in the hot spot (outlined by red dashed lines in Fig. 2). All 4 variants are rare variants in all other ancestries and show little association. One of the top SNPs 9:22093168:A:T (OR=0.89, p= 9.92E-05, and MAF=5%), shows low LD (r2 < 0.2) with the top SNPs from other ancestries, however, is physically close to the top EUR SNP (131 bps away) and the top EAS SNP (9:22092924:A:G) (244 bp away), suggesting the presence of a possible AFR specific signal in the region of association in other ancestries. Neither the homozygous (NAT|NAT) nor heterozygous NAT (NAT|non-NAT-AFR) ancestry groups reached genome-wide significance; however, the heterozygous group showed nominal associations and followed the common trend near 22.1 Mb (Fig. 2).

Across all summary statistics outlined above, we identified 667 genome-wide significant SNPs at the 9p21.3 locus, 298 of which fall within the primary association hotspot (Fig. 1 & 2). The cumulative distribution of minor allele frequencies (MAF) for these significant SNPs reveals broad population differences. Although the homozygous AFR group has lower-frequency variants relative to other groups, fewer than 15% of these variants have a MAF below 10%, suggesting that low-frequency variants and reduced power are unlikely to fully explain the lack of association signals in this group (Fig. S4A). Moreover, this distinction diminishes when the analysis is restricted to variants within the hotspot—the most replicated region at the 9p21.3 locus (Fig. S4B)—where MAF distributions appear more similar across ancestry groups, including homozygous AFR, further underscoring that power differences due to sample size or allele frequency do not account for the observed differences. Lastly, several variants display nearly identical allele frequencies between all groups (EUR, EAS, AMR) yet diverge (Table S3) in their association signals in homozygous African. For example, the lead European SNP (9:22093299:A:T) has comparable MAFs in homozygous Europeans (0.488) and homozygous Africans (0.452), yet shows no significant association in the latter group.

### Cross ancestry population structure at 9p21.3

We analyzed population structure at locus 9p21 using standard metrics, including linkage disequilibrium (LD) and Hudson Fst. To examine the LD structure of CAD associated SNPs in the primary hotspot at locus 9p21 (21.95-22.135 MB), we generated a heatmap plot (R²) for genome-wide significant SNPs that have a MAF > 1% in any of the groups. We observe two LD blocks, with their extent varying by ancestry. EUR exhibits the strongest LD structure, followed by SAS, EAS, and MID (Fig. 1). While the admixed African American group in MVP show some preservation of these LD blocks, they are completely absent in 9p21.3- homozygous AFR individuals; in fact, more than 4 distinct blocks are apparent (Fig. 2). Utilizing AWIgen dataset from continental Africa, we generated regionally specific LD matrices for Eastern, Southern, and Western African regions, and saw strong concordance between homozygous AFR and all three regions(Fig. S2). These findings overall, with a progression of strength of LD consistent with the out of Africa theory of human evolution – weakest in AFR, next in MID, followed by SAS, and then EAS and EUR, suggest variation in LD structure and especially a drastic difference in population structure between AFR and non-AFR ancestries that might be impacting CAD susceptibility at this locus across populations.

To investigate population differences in allele frequencies at the 9p21.3 locus, we calculated the Hudson F_ST_ metric using PLINK2. F_ST_ values reflect average allele frequency divergence across the primary 9p21.3 signal (spanning 21.9 to 22.17 Mb) including all SNPs independent of significance of association. Not all between-group comparisons were feasible because the study datasets were distributed across multiple servers. As expected, the strongest differentiation is observed between African and non-African ancestry groups (Fig. S3A). The largest contrast is between individuals in MVP who are homozygous for the AFR haplotype and those homozygous for the NAT haplotype (F_ST_ = 0.278). A close second is between UKBB Europeans and Western Africans (F_ST_ = 0.2044), followed by UKBB Europeans and Southern Africans (F_ST_ = 0.1959). Eastern African populations exhibit less differentiation and are systematically closer to all other groups compared to their Western and Southern African counterparts, likely due to West Eurasian admixture. Additionally, moderate divergence is observed between East Asian and European groups (F_ST_ = 0.1254). Complementing these results, GSEL(12), a method for detecting enrichment of evolutionary signatures, identified F_ST_ enrichment between Europeans and East Asians, as well as between East Asians and Africans (Fig. S3B), further confirming population structure at this locus.

Overall, these results underscore that genetic drift has occurred at 9p21.3 between African and non-African populations as well as among non-African populations.

### Local heritability of CAD at the locus 9p21.3

We estimated local heritability of CAD at the 9p21.3 locus using BOLT-REML for individual-level data cohorts and LAVA, a summary statistic-based method, for all other instances (Table S1). Local heritability was highest in the Middle Eastern ancestry group (1.4%), likely due to the case–control design of the study, which was heavily enriched for cases. For the other cohorts, local heritability estimates were similar, ranging from 0.1% to 0.2%. In contrast, the AFR homozygous group had an estimate of h² = 0.00013, but with a very large standard error (0.000265), making the result non-significant.

Lastly, to evaluate the contribution of 9p21.3 to PRS performance, we compared inclusion versus exclusion of 9p21.3 variants in the Inouye et al. 2018 metaGRS among EUR and AFR in MVP (Table S26). Overall, exclusion of 9p21.3 resulted in only marginal changes in predictive performance. In AFR individuals, odds ratios (OR ≈ 1.12) and AUC values remained nearly identical (AUC = 0.6005 to 0.6002), whereas EUR individuals showed a modest reduction (AUC = 0.6213 to 0.6186; OR = 1.36 to 1.34). Among AFR individuals, those heterozygous for African ancestry at 9p21.3 showed a slight decrease in AUC (0.5334 to 0.5308), while the homozygous group remained unchanged (OR = 1.11) highlighting the lack of contribution of 9p21 to the PRS predictive value in this group.

### Candidates Variants / Credible Set Identification Using Conditional and Bayesian Fine-Mapping Across Ancestries

Following the association analyses, we performed fine-mapping of the locus 9p21.3 using both conditional analyses and Bayesian methods to better characterize its genetic architecture across all groups. As a sensitivity check, we conducted conditional analyses with both REGENIE (10) and PLINK2 (13) in UKBB EUR. Results were consistent across methods, and for practicality, we used PLINK2 for all subsequent analyses. Conditional analyses in PLINK2 were broadened to SNPs within an expanded 9p21 region (21–23 MB).

In the homozygous EUR in MVP, when conditioning on the top hit (9:22093299:A:T,p=1.19x10^-175^), we observed a secondary signal 9:21816573:G:A at 83 KB from the primary hotspot (Fig. 2A, Table S9). When conditioning on 9:22093299:A:T and 9:21816573:G:A, 9:21964882:CAAAA:C within the primary hotspot showed the strongest association(p = 1.58×10⁻⁵); suggesting a potential presence of a second independent signal at 9p21.3 in homozygous Europeans. In our UKBB EUR conditional analysis this SNP however was not replicated. In BBJ East Asian individuals, conditioning on the top hit (9:22092924:A:G) revealed a secondary cluster of 18 SNPs (lead 9:22114469:G:C, p = 9.58X10-9, AF = 0.61, OR = 1.074, Fig. 3B). The C allele is globally polymorphic, for example frequency 0.51 in EUR. In EAS it is in LD with a 17-SNP haplotype spanning 9:22089264 to 9:22118102 with a frequency of about 0.18. Of the 18 SNPs, six (9:22099568:C:A, 9:22101702:T:C, 9:22102165:C:T, 9:22103341:T:G, 9:22103813:A:G, 9:22114469:G:C) are common and significant in EUR-homozygous, common and nominally significant in Saudi, and common in NAT-homozygous. The remaining twelve are rare in EUR-homozygous and NAT-homozygous, several of which are common in AFR-homozygous, SAS, and Saudi (Tables S8, 20, 21, and 24). In the first conditional pass in BBJ, the risk alleles across this 17-SNP haplotype were not genome-wide significant, and many showed ORs below 1, consistent with negative LD with the A risk allele at the primary SNP 9:22092924 (Table S8). The C allele at 9:22114469 was not a top signal in MVP or UKBB EUR, was in incomplete LD with their lead variants, and was not significant after conditioning. Thus, it is conceivable that it is the unique haplotype of 17 SNPs or some subset of it in the EAS that is conferring independent risk in that population. Furthermore, the section of haplotype of SNPs distal to 9:22114469 does exist in Africans (frequency around 5% - and a sub-haplotype even more frequent) – but none of those SNPs show association in AFR. Thus, the risk conferred by this haplotype may lie in the section proximal to 9:22114469. When conditioning on 9:22092924:A:G and 9:22114469:G:C, the strongest signal was observed at 9:21825075:T:C which is 75 KB away from the primary spot and landed in the secondary signal observed in EUR (Fig. 3B). No secondary signals were observed in other ancestries, including NAT homo/heterozygous, Native Arab, and individuals heterozygous with the African chromosomes (Table S2), although sample sizes were smaller for those groups.

**Figure 3:**
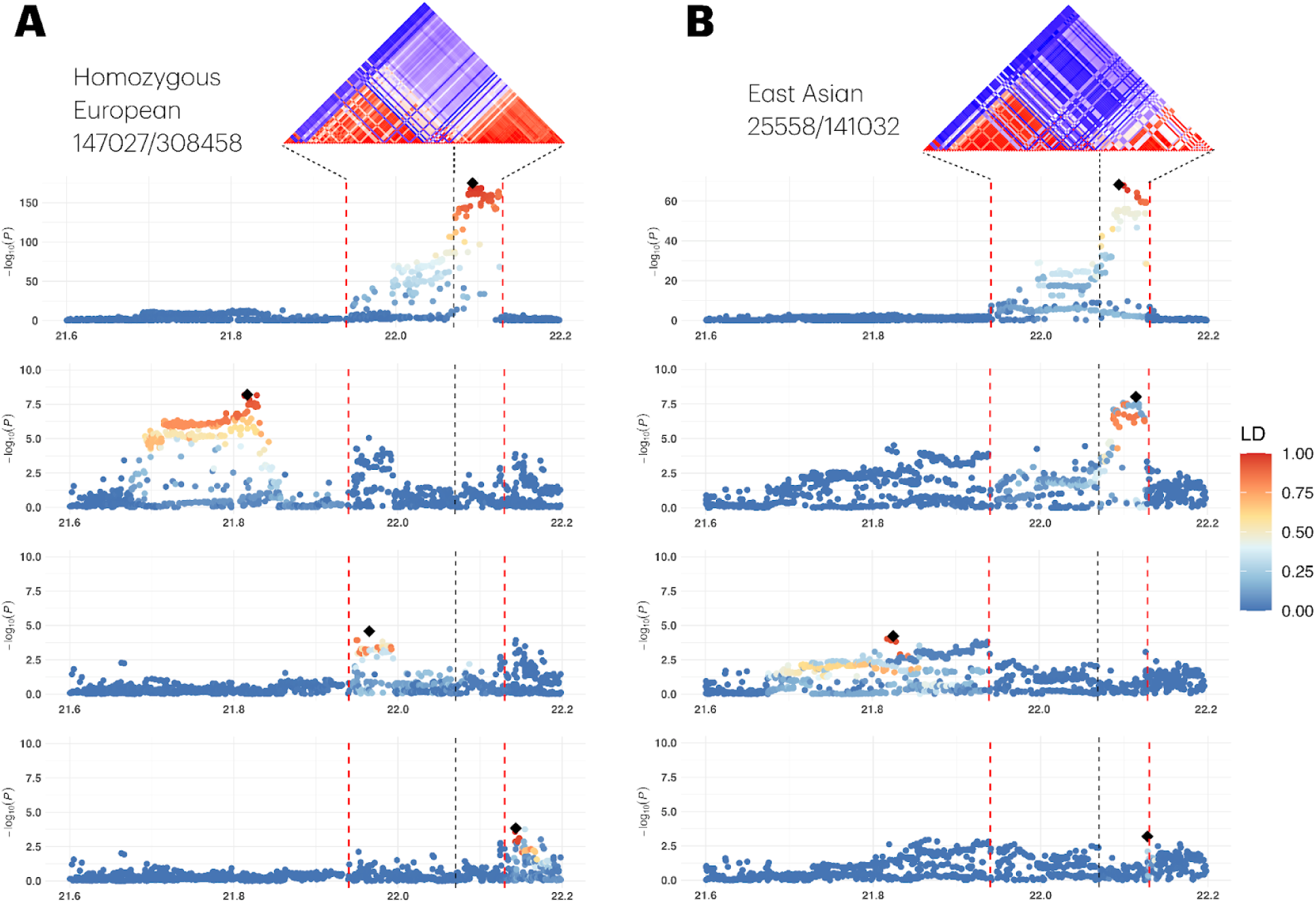
Stepwise conditional analysis at locus 9p21.3 in European and East Asian populations. Results from stepwise conditional analyses conducted separately in (A) MVP individuals homozygous for European local ancestry at 9p21.3 and in (B) East Asian individuals from BioBank Japan (right column). Each column displays a series of locus zoom plots, with the top row showing the marginal association results. Subsequent rows represent association results after conditioning on the lead SNP identified in the preceding step. LD is calculated relative to the top SNP from that specific association run. LD reflects in-sample estimates within each respective cohort.

We also performed Bayesian fine-mapping using SuSiE with summary statistics and in-sample LD matrices, for SNPs with MAF > 5%. In MVP homozygous EUR, five credible sets were identified at 9p21.3—four at the primary hotspot (Fig. S9), three of which containing a single SNP (9:22093299:A:T, 9:21951929:G:GA, and 9:22082375:A:C), and the fifth containing 17 SNPs. Credible set 2, residing in the distal region (21.7–21.83 MB) contains 209 SNPs (Fig. S9, Table S15). However, these fine-mapping results are not fully concordant with our stepwise conditional analysis, and we caution that in very high-LD regions like 9p21.3, SuSiE’s IBSS optimizer can occasionally converge to suboptimal solutions that generate false-positive credible sets as reported by the SusieR authors. Following their suggestions we incorporated their refinement method which did not change the results. In the East Asian BBJ cohort, two credible sets were identified—one containing 4 SNPs and the other containing 18 SNPs (Fig. S9, Table S16). Of the 18 SNPs in the second set, 11 were also significant in the conditional analysis after adjusting for the lead SNP, demonstrating strong concordance between the Bayesian and conditional analyses. Our findings highlight not only the presence of multiple independent signals but also the existence of population-specific candidate variants that may have been missed in prior studies due to differences in LD structure. While LD undoubtedly influences these findings, our results strongly suggest that multiple common causal variants are likely at play at 9p21.3.

### Haplotype Diversity and Its Relationship to CAD Risk at 9p21.3

Given the evidence for multiple candidate causal variants at 9p21.3 and the potential confounding effects of LD, it is important to consider how these variants are organized within broader haplotype backgrounds. Understanding the distribution of these haplotypes and their relationship to CAD risk across diverse populations can provide insight into how ancestry-specific LD patterns may influence both the genetic architecture of this locus and the ability to detect associations within each ancestry and possibly the specific combinations of SNPs directly involved.

Following Tcheandjieu et al. 2022, we expanded the haplotype analysis at the 9p21 locus, using the same six SNPs to construct haplotypes, estimate their frequencies, and assess their association with CAD in East Asian, South Asian, Native Arab, and continental African populations. In addition to replicating the previously reported European results, we found that the haplotypes AACATT and GGTTCA were predominant across all non-African ancestry groups and virtually absent in AFR homozygous in MVP and across all continental Africans groups except East Africans were we observed an increase in frequencies of the haplotypes AACATT (1.7%) and GGTTCA (5.6%) (Fig. S5) likely due to Mideast admixture. Using AACATT as the reference, we observed a consistent CAD risk associated with GGTTCA, except in South Asian and NAT homozygous individuals (Fig. S5) likely due to small sample size. Similarly, the AGTTCA haplotype, which differs from GGTTCA by a single SNP, is also associated with increased CAD risk in several ancestries (Fig. S6).

We further expanded the haplotype analysis to a larger set of SNPs. When linkage disequilibrium is low, each additional SNP substantially increases the number of possible haplotypes, making exhaustive analysis computationally infeasible. To address this issue, we selected a subset of significant SNPs at 9p21.3 and constructed a theoretical haplotype composed entirely of the protective alleles. For each individual, we computed the Hamming distance—the count of mismatched alleles—between this theoretical haplotype and their observed haplotype (Fig. S12). We also took the sum of hap1 and hap2 which resulted in a total Hamming distance that reflects the number of risk alleles carried by each individual (Fig. 4A). Among African ancestry groups—including homozygous AFR, Eastern Africa, Western Africa, Southern Africa —we observe a unimodal, bell-shaped distribution centered around intermediate distances (Fig. 4A). Interestingly, concordant with the 6-SNP haplotype analysis, Eastern African individuals show haplotypes containing a greater number of risk alleles when hap1 and hap2 are examined separately (Fig. S12). This pattern is reflective of haplotype diversity, resulting in many individuals carrying haplotypes that are divergent from both the full protective and risk configurations.

**Figure 4:**
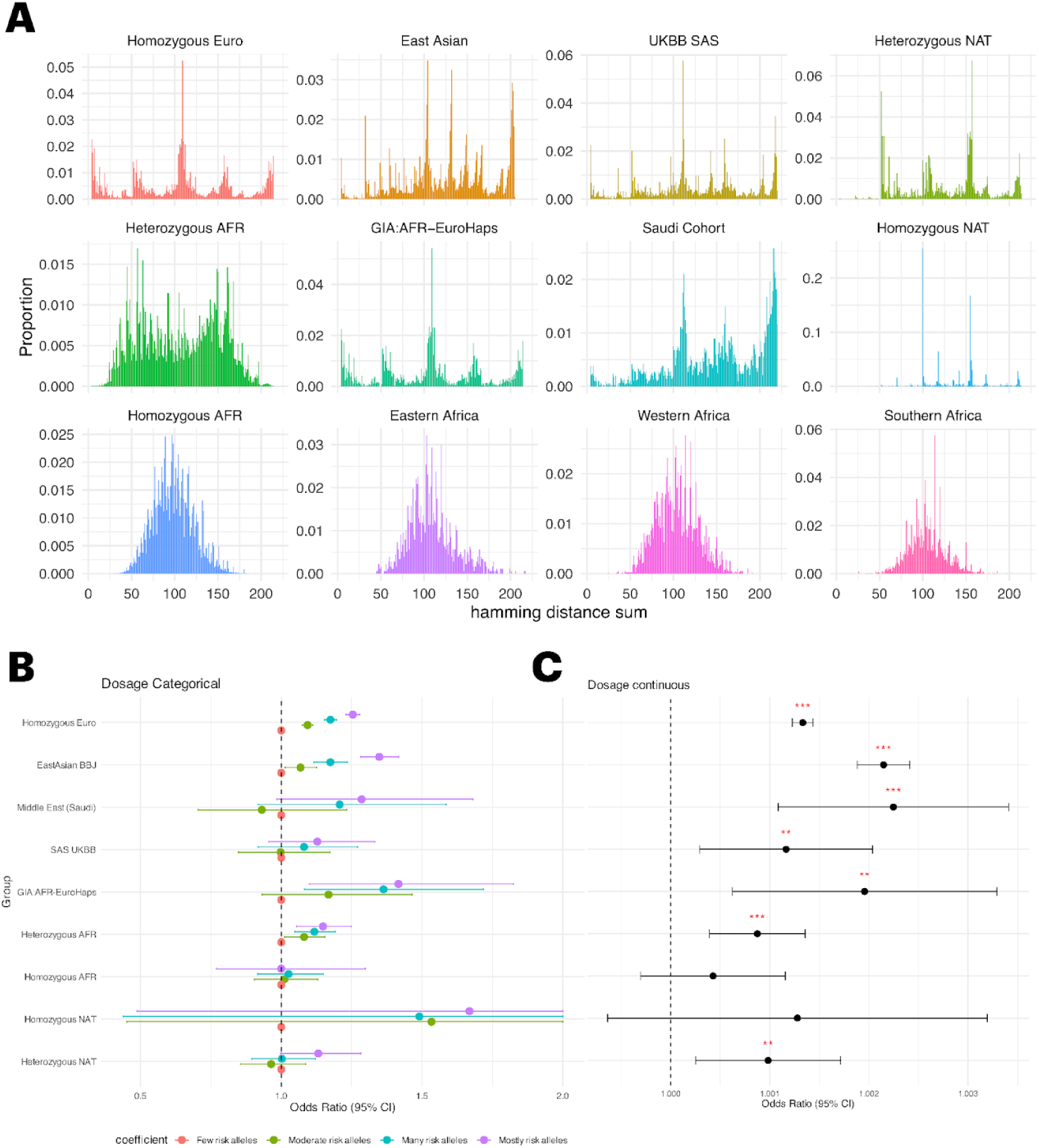
Hamming distance across cohorts at locus 9p21.3. Hamming distances were calculated between each individual’s haplotypes, composed of common CAD-associated SNPs, and a theoretical reference haplotype carrying all non-effect alleles (NAH). Distances were computed separately for each haplotype (hap1 and hap2), and the combined Hamming distance (hap1 + hap2) was used in this analysis. Distributions of hap1 and hap2 distances are provided in the Supplement. **A.** Distribution of total Hamming distances across all studied ancestry groups. **B.** Association between Hamming distance and CAD. Individuals were grouped into four Hamming distance categories—“few risk alleles,” “moderate risk alleles,” “many risk alleles,” and “mostly risk alleles”—by dividing the full range of summed Hamming distances into four equal-width intervals. Association testing used the “few risk alleles” group as the reference category. Sex and PCs 1-5 were used as covariates. **C.** Association between Hamming distance and CAD. Here hamming distance was coded as a continuous trait.

The African heterozygous group shows a more bimodal distribution, reflecting a mixture of haplotypes: one mode likely representing chromosomes more similar to the protective configuration, and another mode aligned with risk-associated or highly divergent haplotypes (Fig. 4A). Non-African groups—particularly EUR and SAS, display highly multimodal or spiked distributions, indicating the presence of dominant haplotype configurations with relatively uniform divergence from the theoretical protective sequence. The strong peaks at specific Hamming distances suggest less recombination and greater haplotype homogeneity in these populations (Fig. 4A, S12). Furthermore, individuals classified as AFR, who are homozygous EUR at 9p21.3 share similar distribution with homozygous EUR; underscoring the impact of admixture in generating distinct haplotype classes within individuals (Fig. 4A). The Hamming distance distribution in EAS BBJ and Saudi cohort are noticeably left-skewed.

Using Hamming distances (equivalent to the total number of risk alleles), we assessed association with CAD. When coded as a continuous variable (Fig. 4C), higher divergence from the protective haplotype is associated with increased risk in Homozygous EUR, Heterozygous AFR, GIA:AFR-EuroHomozygous, Heterozygous NAT, South Asian UKBB, BBJ EAS, and the Middle East (Saudi) cohort. Positive trends were observed in homozygous NAT and AFR groups; however, these did not reach statistical significance owing to wide confidence intervals. Notably, the point estimate for homozygous AFR was the smallest across all groups.

To further investigate the relationship between haplotype divergence and CAD risk, we categorized total Hamming distances into four bins—Few, Moderate, Many, and Mostly risk alleles—with the “Few risk alleles” group serving as the reference (Fig. 4B, Table S17). Across multiple groups, we observed a clear dose-response pattern: odds ratios increased progressively with higher levels of divergence from the protective haplotype. This trend is especially pronounced in Homozygous EUR, EAS, Heterozygous AFR, and GIA:AFR-EuroHomozygous, where individuals with “Mostly risk alleles” showed the highest CAD risk.

### Pleiotropic Assessment of 9p21.3 Across Local Ancestry Groups

Because the 9p21.3 locus is highly pleiotropic, with previous studies reporting associations to multiple traits in non-African ancestries, we next investigated whether similar patterns emerge in African and non-African ancestry groups in MVP. We performed a phenome wide association study (PheWAS) in MVP within each local ancestry group. To increase sample size for NAT, we combined heterozygous and homozygous NAT individuals. When comparing p-values between AFR homozygous and EUR homozygous, we observed little correspondence (Fig. 5). In contrast, despite the smaller sample size, AFR heterozygotes showed a stronger trend with the EUR homozygous group, consistent with shared EUR haplotypes contributing to more similar association profiles (Fig. 5). We also observed a similar trend in NAT PheWAS when compared to EUR. As expected, type 2 diabetes and related phenotypes replicated in both the African heterozygous and NAT groups, but were not observed in the African homozygous group. Interestingly, in the AFR homozygous group we identified a SNP in the primary hotspot associated with the phecode “Hypertrophic obstructive cardiomyopathy” (HOC) (9:22015111:T:C, OR = 1.9, p = 9.27 × 10⁻⁸). In addition, 8 other SNPs in LD with 9:22015111:T:C showed suggestive signal (p < 3e-5) for the same phecode (9:22024339:G:A, 9:22058819:C:T, 9:21997752:A:C, 9:21997723:T:C, 9:22013815:A:G, 9:22015782:C:T, 9:22033362:C:A, 9:22033393:A:C). Notably, all 9 variants are rare (< .7%) in both NAT and EUR MVP groups. Furthermore, we constructed LD heatmaps for our various groups consisting of all phenotype associated SNPs in EURO homozygous (Fig. S11). Similar to our CAD based LD heatmaps in Fig. 1 & 2, we see the most broken down LD for AFR homozygous.

**Figure 5:**
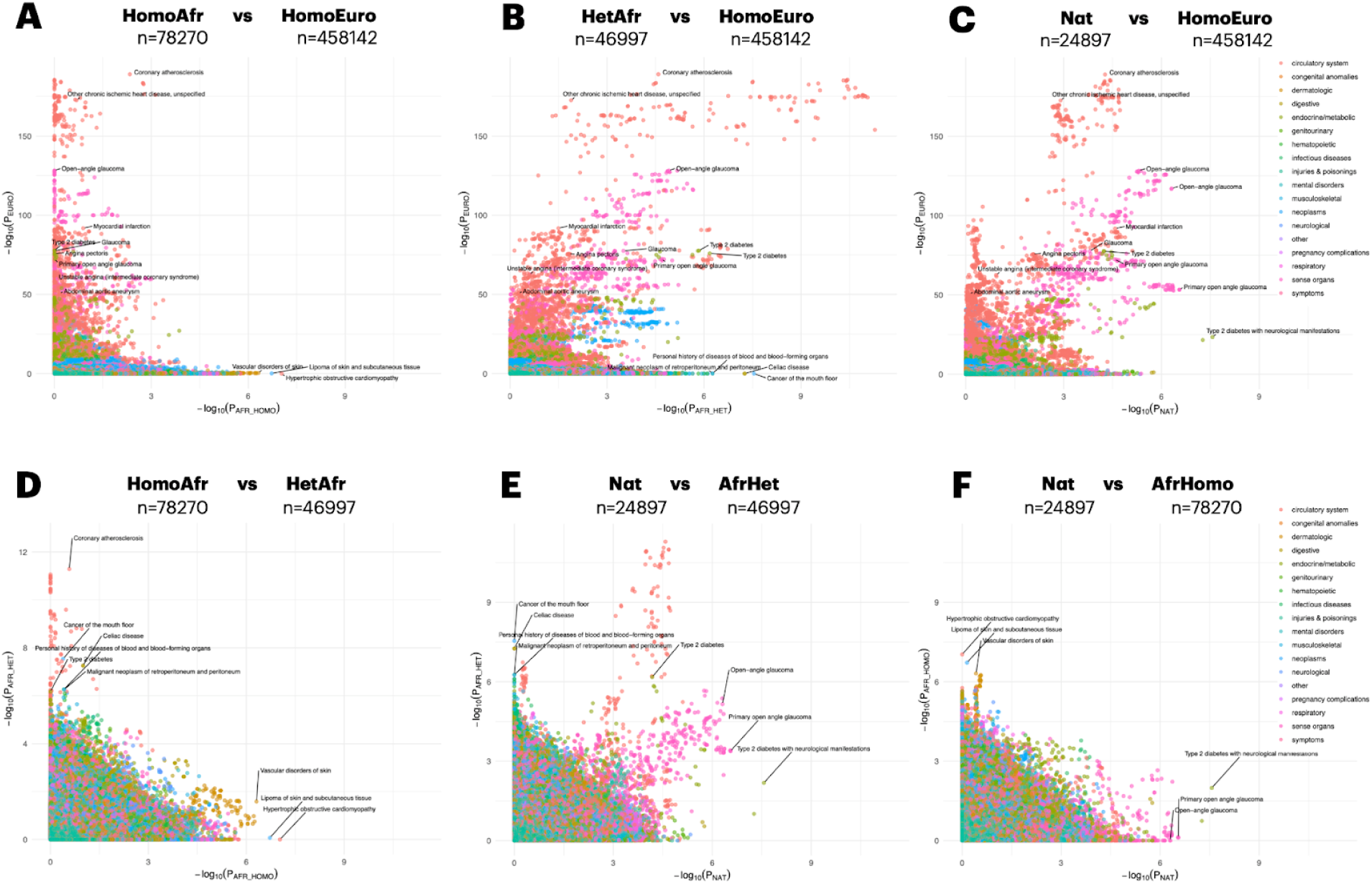
Comparison of phenome-wide association results across local ancestry groups in MVP. Phenome-wide association (PheWAS) was conducted using phecodes in MVP individuals stratified by local ancestry at 9p21.3, focusing on homozygous and heterozygous ancestry groups for African, European, and Indigenous American chromosomes. NAT group includes both heterozygous and homozygous individuals in order to increase sample size. Panels (A-F) shows a pairwise comparison of −log₁₀(p-values) between two groups. Variants with minor allele frequency <5% or absolute log odds ratio >2 were excluded. Phenotypes with fewer than 300 samples were also removed. Points are colored by phenotype category (system), and selected points are labeled if they reached genome-wide significance in the European group or nominal significance in other groups. To reduce clutter, only the top SNP for each phecode was selected for labeling. Full association details are provided in the Methods section.

A complementary PheWAS using available laboratory measurements in MVP revealed a similar trend to the phecode-based analysis (Fig. S10). Notably, SNPs associated with platelet and monocyte levels appeared in the African homozygous group and replicated in Europeans, whereas previously reported blood traits such as HbA1c and glucose were associated only in the EUR homozygous and AFR heterozygous groups. These results suggest that the difference in population structure at 9p21.3 between AFR and non-AFR ancestry individuals impact susceptibility to diseases previously reported at the locus beyond CAD.

## Discussion

In this study, we adopted an ancestry-guided approach to evaluate one of the most significant risk loci for CAD discovered by GWAS. Rather than obscuring population differences through multi-ancestry analyses, we stratified our analyses by ancestry to better characterize the genetic architecture of this locus. Our examination of the 9p21.3 region across multiple ancestries highlights both its complexity and the critical role of MAF and LD in shaping association signals. While this locus is among the most reproducible CAD associations in EUR, EAS, and other non-African populations, we observed no replication of the same variants in African ancestry individuals – even within the large, well-powered MVP cohort. Notably, this lack of replication cannot be attributed simply to lower allele frequencies or reduced power: many CAD-associated SNPs associated in EUR and other non-African populations display polymorphic frequencies in African populations, yet have null association signals. It is important to note that we observe population differentiation in our Fst analysis which took into account an expanded set of SNPs at the locus 9p21.3 hotspot beyond just CAD significant SNPs. Both levels of evidence, population-wide divergence and trait-specific allele frequency distributions, should be considered in interpreting replication gaps.

### LD STRUCTURE, HAPLOTYPE DIVERSITY, AND RISK ALLELE DISPERSION PLAY PIVOTAL ROLES IN LACK OF 9p21.3 CAD REPLICATION

LD is a well-established factor in association studies, often discussed in two key contexts. First, LD enhances the ability to detect associations by allowing non-causal SNPs to tag nearby causal variants. Second, that same LD complicates fine-mapping by making it difficult to distinguish among tightly correlated candidates. Recent high-throughput functional studies have begun to disentangle this problem by demonstrating that many complex disease loci harbor causal variants within the same region (14). LD also shapes haplotype diversity: lower LD increases the number and variability of haplotypes, and it has been shown that haplotype diversity, just allelic diversity, decreases with distance from Africa, consistent with human demographic history (15).

Our analyses suggest that LD structure plays a pivotal role in the patterns of 9p21.3 CAD signal (non)replication and population differentiation that we observed. In non-African populations, CAD-associated SNPs reside in extended LD blocks that can efficiently carry all risk alleles. In contrast, LD is substantially weaker in homozygous African ancestry at 9p21.3, breaking apart multi-SNP haplotypes and dispersing risk alleles across a broader set of configurations. This is also true for EAS and AMR, SAS and MID but to a much lesser extent. This is further supported by our conditional results, which identified independent association signals, including population-specific credible sets that disappear or emerge because of LD structure.

The haplotype analyses reinforce this interpretation. Across non-African populations, we observed a small number of dominant haplotype configurations—often carrying multiple risk alleles—that are rare or absent in African ancestry. African heterozygotes show haplotype distributions and CAD associations more similar to European homozygotes, consistent with the presence of shared European-derived haplotypes and dominant (heterozygous) expression. In the homozygous African group, however, greater haplotype diversity appears to dilute the cumulative signal of multiple causal alleles, attenuating association signals even when individual allele frequencies are similar. Our Hamming distance approach further demonstrated that CAD risk increases with greater divergence from a theoretical protective haplotype.

### LACK OF REPLICATION LOCUS 9p21.3 ASSOCIATION ACROSS TRAITS IN NON-ADMIXED AFRICAN GROUPS

The PheWAS analysis extends our LD hypothesis beyond CAD. While EUR homozygotes, AFR heterozygotes, and NAT (homozygous + heterozygous) share trait associations such as type 2 diabetes and glycemic traits, these signals are absent in homozygous African ancestry despite comparable allele frequencies for some lead variants. These findings suggest that the same mechanisms underlying the lack of CAD association in AFR homozygotes, namely weaker LD, greater haplotype diversity, and dispersed risk alleles, also extend to other phenotypes at this locus. In the AFR homozygous group, only certain continuous traits (e.g., platelet count, monocyte count) replicate, likely reflecting the higher statistical power of quantitative phenotypes, or the high enrichment of genetic signal for blood cell count in AFR ancestry (16,17). It is important to note that in the AFR homozygous group we observed novel SNP associations with CAD (9:22093168:A:T, OR = 0.89, p = 9.92 × 10⁻⁵), hypertrophic obstructive cardiomyopathy (HOC) (9:22015111:T:C, OR = 1.9, p = 9.27 × 10⁻⁸), and glucose levels (9:22325667:C:CT, b = –0.86, p = 8.54 × 10⁻⁷), all of which are rare (<1%) in other ancestries. While none of these variants reached genome-wide significance; their locations overlap with previously reported association regions, suggesting potential ancestry-specific signals that warrant further investigation. For example, the glucose lead SNP lies within a locus previously implicated in type 2 diabetes risk. Importantly, the novel AFR-specific signals we observe do not contradict our model of multiple causal SNPs situated on haplotypes that are broken down by LD in homozygous African populations, but rather illustrate how distinct associations may emerge when these haplotypes are decomposed.

### LOCUS 9p21.3 REGULATORY MECHANISMS SUGGEST A MULTIPLE CAUSAL SNP ARCHITECTURE

The 9p21.3 locus contains a compact cluster of genes—*CDKN2A, CDKN2B, MTAP,* and *ANRIL* (*CDKN2B-AS1*)—that regulate key cellular programs relevant to plaque biology. Converging evidence suggests that CAD risk alleles at 9p21.3 impact *ANRIL* expression, splicing, and linear/circular isoform ratios, making *ANRIL* a likely mediator of locus effects, while not excluding additional contributions from the other 9p21.3 genes (5,18). Mechanistically, *ANRIL* acts in cis by recruiting PRC1/PRC2 to enforce H3K27me3-mediated repression at *CDKN2A*/*CDKN2B* (5,19), and is hypothesized in trans to act via linear-isoform Alu-guided RNA–DNA contacts at distal loci (5,20).

In iPSC-derived VSMCs, deleting a 60-kb block spanning the European risk vs non-risk haplotypes produced clear transcriptional changes in many CAD loci and lowered upstream *ANRIL* when the risk block (*ANRIL* exons 10–19) was removed, consistent with enhancer-driven upregulation on the risk background (6). Overexpressing ANRIL in non-risk knockout cells recapitulated several—but not all—risk phenotypes, suggesting that additional elements within 9p21.3 contribute. A follow-up single-cell study showed that risk-haplotype VSMCs diverge toward osteogenic and chondrogenic trajectories; deleting the risk haplotype mitigated the pro-calcific phenotype, and overexpression of a short ANRIL isoform in non-risk VSMCs partially reproduced these transcriptional shifts (7). Complementing these perturbations, a vascular splicing QTL (rs10217586) colocalizes with CAD and is common across ancestries (MAF ≈ 0.46), yet associates in non-African groups and not in non-admixed Africans—consistent with LD-context dependence and a distributed, modest-effect architecture on *ANRIL* regulation and splicing (21).

Reinforcing this view, an enhancer-perturbation screen in human coronary SMCs mapped a multi-enhancer regulatory surface at 9p21.3—linking ten enhancers to CDKN2A/CDKN2B/MTAP—and found that phenotypic shifts occurred only when multiple enhancers were co-activated, supporting additive/interactive modest effects rather than a single causal variant (22). Follow-up CRISPRa showed *ANRIL* was upregulated only when all ten enhancers were activated. The authors also nominated rs1537372, which disrupts an AHR/ARNT site in one enhancer; its risk allele is ∼4% in YRI and was proposed to contribute to attenuated African signals. However, in our homozygous-African analysis rs1537372 showed no effect (OR = 0.983, p = 0.56) (22).

Taken together, evidence supports a polymechanistic 9p21.3 with an *ANRIL*-centric hub: modest cis variants likely tune *ANRIL* expression, splicing, and linear/circular balance in VSMCs—consistent with lncRNA biology (weak exon conservation, constraint at promoters/splice sites/short motifs). These functional observations, when viewed in the context of our ancestry-stratified analyses, illustrate how demographic history and LD structure could shape locus behavior across populations.

### THE STORY LOCUS 9p21.3 STORY GENERALIZED ACROSS THE GENOME

Our results suggest that the long-standing lack of replication for 9p21.3 CAD associations in African-ancestry populations is driven by greater haplotype diversity that attenuates the signal of individual variants. Moreover, this phenomenon is unlikely to be unique to 9p21.3: for many complex-trait loci with multiple causal variants (23), the local configuration of variants is shaped by ancestry-specific LD. Future work will require systematic evaluation of trait loci across the genome. Addressing this detection bias from dispersed causal SNPs will require new methodology and continued growth in African-ancestry sample sizes.

Gaining insight into 9p21.3 requires an ancestry-specific strategy. Multi-ancestry GWAS boost power (24,25), but can blur population-specific signals at complex loci; analyzing ancestries separately—and exploiting their differences—better reveals architecture where several small-effect alleles act together. Our findings suggest that fine-mapping at complex regulatory loci such as 9p21.3 is unlikely to resolve to a single causal variant (e.g., a rare founder variant with additional rare alleles in the same regulatory elements) but instead often involves multiple causal SNPs whose distribution is shaped by local LD patterns and allele frequencies. Leveraging cross-population differences in MAF and LD enables finer mapping of common causal variants, characterization of haplotype diversity, and explanations for signals missed in specific groups. This locus illustrates how genetic diversity can illuminate disease biology that remains hidden in homogeneous cohorts.

### STRENGTHS AND LIMITATIONS

This study represents the first cross-ancestry fine-mapping of a major CAD locus that includes European, African, East Asian, South Asian, Middle Eastern, and Admixed American populations with Indigenous American ancestry. We performed fine-mapping using large sample sizes for African, European, and East Asian groups. East Asian ancestry currently represents the second-largest cohort after Europeans, and its inclusion was critical for characterizing cross-ancestry patterns at 9p21.3.

A primary limitation of this work is the comparatively small sample sizes available for Middle Eastern, South Asian, and Indigenous American groups. Expanding representation in these populations will be essential for uncovering additional ancestry-specific variants and refining the locus’s multi-causal architecture through conditional and joint analyses.

Finally, the validity of our working hypothesis is not contingent upon how present-day 9p21.3 haplotypes arose; however, important evolutionary questions remain. Do the observed haplotype spectra primarily reflect stochastic forces—genetic drift and bottlenecks associated with Out-of-Africa demography—or have evolutionary pressures such as local adaptation and background selection also contributed to shaping this region

## Conclusion

In this study, we dissect the association between CAD and variants at the locus 9p21.3 across multiple ancestries. We showed that the lack of 9p21.3 replication in African ancestry reflects weaker LD and greater haplotype diversity, which distribute risk alleles across configurations and weaken the statistical signal while, in other ancestries, risk alleles are consolidated on fewer haplotypes, thus strengthening the signal. The complex functional landscape and haplotype architecture at 9p21.3 together accommodate ancestry-specific signals, such as the described East-Asian conditional hits, and allow for local cis-epistasis among *ANRIL*-linked enhancers and splicing elements that collectively modulate locus-wide regulatory output. Overall, it is likely that the demographic history at this locus—including the Out-of-Africa bottleneck and subsequent founder events—tightened LD in non-African populations, co-packaging functional alleles on shared risk haplotypes, whereas many African haplotypes separate these elements, potentially diluting aggregate effects and further attenuating association strength.

## Methods

### Cohorts and Populations Studied

Our analysis integrates multiple datasets to comprehensively capture the genetic diversity of the 9p21 locus. Specifically, we leverage data from the UK Biobank (UKBB)(26), Biobank Japan (BBJ) (27), Million Veteran Program (MVP) (28,29), H3Africa/AWI-Gen (30), and a Saudi cohort from the King Faisal Specialist Hospital and Research Centre (KFSHRC)(31) Collectively, our study includes individuals from a wide range of ancestral backgrounds, including European (MVP, UKBB), East Asian (BBJ), Middle Eastern (KFSHRC), South Asian (UKBB), Admixed American (MVP), African American (MVP), and continental African populations spanning Western, Eastern, Central, and Southern Africa from H3Africa/AWI-Gen cohort. European, Admixed American, and African American groups in MVP were further decomposed into local ancestries described below. Detailed sample sizes for each cohort are provided in supplementary table S0.

Coronary artery disease (CAD) phenotypes were available in all cohorts except for H3Africa/AWI-Gen.

### CAD Phenotype

For each cohort, CAD phenotypes were derived from EHRs as previously described in (2–4). For each cohort, phenotype data integrate inpatient and outpatient International Classification of Diseases (ICD 9/10) diagnosis and procedure codes, Current Procedural Terminology (CPT) procedure codes, clinical laboratory measurements, and medications. Report of diagnostic imaging or actual imaging is available for some cohorts such as the UKBB and the MVP. Coronary artery diseases was be defined by: The presence of 1 or more of ICD 9 or 10 codes (410, 410.0, 410.00-02, 410.1, 410.10-12, 410.2, 410.20-22, 410.3, 410.30-33, 410.4, 410.40-42, 410.5, 410.50-52, 410.6, 410.60-62, 410.7, 410.70-72, 410.8, 410.80-82, 410.9, 410.90-92, 411.0, 411.1, 411.81, 411.89, 412, 414.00, 414.01-05, 414.2-4, 414.8, 414.9, V45.81-82, I21, I21.0, I21.01-02, I21.09, I21.1, I21.11, I21.19, I21.2, I21.21, I21.29, I21.3 4, I21.9, I21.A, I21.A1, I21.A9, I22, I22.0-2, I22.8, I22.9, I23, I23.0, I23.1-8, I24, I24.0-1, I24.8-9, I25.1-2, I25.5-6, I25.70-73, I25.79, I25.810, I25.82-84, I25.89, I25.9, Z95.1, Z98.61, I20.0) as an outpatient or an inpatient and/or hospital discharge for coronary artery revascularization procedure.

### Genomic data

For our analysis, we utilized previously-imputed genotype data provided by the UK Biobank (study accession #87255), Biobank Japan, and the Million Veteran Program (27–29). In H3Africa/AWI-Gen and Saudi cohorts, genotype imputation was performed using the Michigan Imputation Server with the 1000 Genomes Phase 3 v5 reference panel (32). The imputation process involved two key steps: first, haplotype phasing is conducted using Eagle, which efficiently reconstructs haplotypes from genotype data; second, missing genotypes were imputed using Minimac4 (32). Additionally, we leveraged the Whole Genome Sequence data from the UK Biobank (study accession #87255) for common and rare variant analysis.

All analyses were conducted using human genome build 37, except for the UK Biobank whole-genome sequencing (WGS) data which used build38 in our rare variant analysis.

### Ancestry Inference

#### Global Ancestry Inference

Global ancestry groups were used in both UK Biobank and Million Veteran Program. The Saudi cohort as well as the AWIGen dataset did not require additional ancestry inference for populations, as this information is provided in the available meta data for each of these cohorts. Ancestry inference in MVP was performed using a genetically inferred ancestry approach (GIA) (33). Genetically inferred groups in MVP included European, African American, and Admixed American (considered to be largely self-reported Hispanic/Latino with Indigenous American ancestry) groups. Global ancestry inference in the UK Biobank (UKBB) was performed using a Random Forest (RF) classifier, as follows. Principal component analysis (PCA) was first conducted on the 1000 Genomes Project (1000G) and Human Genome Diversity Project (HGDP) reference panels, retaining the first 10 principal components (PCs) for downstream analysis. A Random Forest classifier was then trained using super population labels from 1000G/HGDP within this PCA space. UKBB individuals were projected onto the same PCA space and classified into EUR, AFR, SAS, EAS, AMR, and MID (Middle East) using the trained model. We refer to these groups as genetically inferred ancestry (GIA) groups which roughly correspond to self-identified race/ethnicity groups .

#### Local Ancestry Inference

Local ancestry inference (LAI) in the Million Veteran Program (MVP) was performed using Gnomix (34), using non-admixed individuals from the 1000G and HGDP as the reference training panel. For training, we selected non-admixed individuals identified in the original Gnomix study(34). The final training sample set included 352 AFR, 352 EAS, 153 EUR, 88 NAT, and 176 SAS. The final training SNP set included 335,870 SNPs on chromosome 9, of which 2,022 SNPs fell within the regions used in our LAI analysis.

Admixed chromosomes were decomposed into five continental components: African (AFR), European (EUR), East Asian (EAS), South Asian (SAS), and Indigenous American (NAT). The resulting 9p21.3 locus spanned 5 distinct but contiguous LAI intervals (21,650,660–21,748,433; 21,748,434–21,912,417; 21,912,418–22,134,172; 22,134,173–22,361,961; and 22,361,962–22,470,474). For each interval, we assigned each individual’s chromosomes to EUR, AFR, EAS, SAS, or NAT using the highest-probability local-ancestry call. We then defined heterozygous (e.g., AFR|non-AFR) and homozygous (e.g., AFR|AFR) ancestry groups per interval. In the heterozygous NAT group, AFR chromosomes were also excluded.

#### Association and Conditional Analysis

Standard association analyses were first performed using PLINK2(13) across all cohorts with available coronary artery disease (CAD) phenotypes (UKBB, MVP, BBJ, Saudi Cohort). Sex, age, and the first eight principal components were included as covariates for all association analyses. The following filters were included in our analyses: MAF > .1%, imputation mach-r2 of .4 or greater, and minor allele count (mac) > 10. Since PLINK2 does not account for relatedness as linear mixed model (LMM) methods do, related individuals were excluded from conditional analyses. Kinship estimation was performed using KING software, and second-degree and closer relatives (i.e. estimated kinship > 0.125) were removed across all cohorts to minimize confounding and overestimating statistical significance due to genetic relatedness. To validate PLINK2 results and ensure robustness against population structure and relatedness, REGENIE(10) was additionally applied using the same model and filters in the UKBB European ancestry group.

Conditional analyses were also conducted using PLINK2. A minor allele frequency (MAF) threshold of 5% was applied in the conditional analysis to restrict the analysis to common variants, ensuring sufficient statistical power and reducing potential biases from rare variant associations that may be driven by limited sample sizes. After each association test, the most significant SNP (lowest p-value) was identified and iteratively added to the conditioning list for subsequent analyses. All conditional analyses assumed an additive genetic model. In addition to conditional analyses, we conducted Bayesian fine mapping using the susie_rss function in the SusieR package in R (35). In-sample LD matrices were used with their accompanying summary statistics.

To examine population-specific genetic architectures, analyses were stratified by ancestry at multiple levels. First, GIA (see Global Ancestry Inference section)—European, African, East Asian, South Asian, and Admixed American— were analyzed to capture association signals within more admixed continental groupings.

Second, leveraging local ancestry inference, homozygous and heterozygous ancestry groups for the major ancestral populations were separately identified for each of the 5 LAI intervals at 9p21 (see Local Ancestry Inference section). For conditional analysis, groups were defined based on three contiguous intervals (21,748,434–21,912,417; 21,912,418–22,134,172; 22,134,173–22,361,961). Homozygous groups included individuals who were homozygous across all three intervals at 9p21.3, whereas heterozygous groups consisted of individuals carrying at least one copy of the respective ancestry chromosome but not homozygous across all three intervals.

### Rare Variant Analysis

Rare variant analysis was conducted in the UKBB using whole-genome sequencing (WGS) data. REGENIE(10) was used for both single-variant association testing and burden analysis. Sex, age, and the first 8 principal components were included as covariates. Due to insufficient case-control numbers across most ancestry groups, rare variant analysis was limited to individuals of GIA European ancestry (see Global Ancestry Inference section). Variants with minor allele counts < 20 were excluded from the analysis.

### Population structure (Fst, LD, Evolutionary Signatures)

To quantify genetic differentiation between local ancestry groups, we calculated Fst using PLINK2 with Hudson’s estimator, restricting analyses to cohorts that were co-located on the same server. Linkage disequilibrium (LD) for each ancestry group was estimated using PLINK2 with the `--r2` flag.

We assessed signatures of natural selection by running GSEL (12) on GIA EURO summary statistics using the tool’s default settings. GSEL LD-clumps trait loci and tests enrichment/depletion of predefined evolutionary annotations by comparing observed values to frequency/LD-matched backgrounds (using 1000G and SNPSnap database specific to each ancestry), returning trait-level enrichment summaries and locus-level z-scores/empirical p-values.

### Local Heritability Analysis

We estimated locus-specific SNP-heritability (h²_local) for CAD within a fixed window on chromosome 9 spanning 21,000,000–23,000,000 bp, chosen to encompass the full range of significant SNPs identified in our analyses. For South Asian and East Asian groups, where individual-level genotypes were not available, we applied LAVA (36) in univariate mode to GWAS summary statistics, restricting to biallelic variants with MAF ≥ 1% and using ancestry-matched LD reference panels.

For cohorts with individual-level data, we used BOLT-REML(37) to perform regional REML on the same chr9:21.0–23.0 Mb window (again limited to MAF ≥ 1%), including sex, age, and PCs 1–8 as covariates. This yielded locus-level variance component estimates (h²_local) with standard errors on the observed scale.

### Haplotype Analysis

Haplotype analysis was conducted following the approach of Tcheandjieu et al. 2020. Specifically, SNPs that reached genome-wide significance (P < 5 × 10⁻⁸) in the White/European participant meta-analysis were selected and further filtered based on a minor allele frequency (MAF) > 10% and linkage equilibrium (r² < 0.05) in homozygous African participants, resulting in a final set of six independent SNPs. These six SNPs were then used to perform a haplotype trend regression within this region, utilizing the expectation–maximization algorithm as implemented in the R package haplo.stats.

### Hamming Distance Analysis

Using summary statistics we generated for 9p21.3, a comprehensive SNP set was generated based on the following criteria: (1) genome-wide significance in at least one ancestry group, (2) fall within the primary 9p21.3 hotspot between 21.9 MB and 22.17 MB. (3) minor allele frequency (MAF) ≥ 5% across all included ancestries, and (4) phased status in all studied cohorts. This selection resulted in a set of 111 SNPs.

An all-non-risk haplotype was then constructed, representing a haplotype that exclusively contains the protective alleles (Beta < 0) in European summary statistics. Individual-specific observed haplotypes were generated, and the Hamming distance—the number of differing alleles between each observed haplotype and the all non risk haplotype—was calculated for both haplotypes for each participant. Greater distance from the all non risk haplotype represents haplotypes that carry more risk alleles. Hamming distance was subsequently included as a covariate in a generalized linear model (GLM) to assess its relationship with CAD both as a continuous and categorical variable. For the categorical analysis, hamming distance was recoded into an four-level factor using R’s cut() across the observed range, with levels “Few risk alleles,” “Moderate,” “Many,” and “Mostly risk alleles.” In the GLM, we used “Few risk alleles” as the reference, so each category’s coefficient estimates the log-odds contrast relative to that group.

### Phewas in Million Veteran Program

PheWAS was conducted in MVP using both previously defined phecode-based phenotypes and available laboratory measurements (phelabs). Sex-specific phecodes were excluded. For phelabs, all laboratory traits included maximum, minimum, mean, and median values; in our analyses we focused on the median value per individual.

Association tests were performed separately for all phecodes using PLINK2, adjusting for sex, age, and the first eight principal components. Second-degree and closer relatives were removed. Similar to the CAD associations, we leveraged the local ancestry inference calls, and homozygous and heterozygous ancestry groups for the major ancestral populations were separately identified for each of the 5 LAI intervals at 9p21.3.

### PRS analysis

Polygenic risk scores were computed in AFR and EUR participants from the Million Veteran Program using the metaGRS weights described in Inouye et al. 2018 (38). Two versions of the score were generated: (1) the full metaGRS including all genome-wide weights, and (2) a modified score excluding variants within the 9p21.3 locus. Within each ancestry group, individuals were further stratified by their local ancestry at 9p21.3.

Logistic regression models were fitted with coronary artery disease (CAD) as the outcome under three specifications: (i) CAD ∼ PRS, (ii) CAD ∼ sex + age + PC1–PC10, and (iii) CAD ∼ PRS + sex + age + PC1–PC10. Model performance was assessed using the AUC curve.

## Data Availability

All data produced in the present work are contained in the manuscript

## Acknowledgment

We gratefully acknowledge the Veterans who participated in the VA’s Million Veteran Program. This research is based on data from the Million Veteran Program, Office of Research and Development, and Veterans Health Administration, with support from MVP000, VA Merit Award #I01-BX003362 (Chang/Tsao) and the Department of Veterans Affairs (VA) Informatics and Computing Infrastructure (VINCI), including data analytics conducted by its Precision Medicine research team, which is funded under the research priority to Put VA Data to Work for Veterans (VA ORD 24-D4V-02). This work was also supported by the American Heart Association Predoctoral Fellowship (Award ID: 24PRE1201199) and the Lamond Fellowship. . This publication does not represent the views of the Department of Veterans Affairs or the United States Government.

## Author contributions

H.A. conducted analyses, S.K. conducted analyses in BBJ. H.A. and C.T. wrote the manuscript, with input and revisions provided by N.R., S.C., F.A., A.H., K.I., T.A..

### Abbreviations

CAD: Coronary Artery Disease
GIA: Genetically Inferred Ancestry
LAI: Local Ancestry Inference
LD: Linkage Disequilibrium
MVP: Million Veteran Program
WGS: Whole Genome Sequencing

---

**Supplementary Figure 1:**
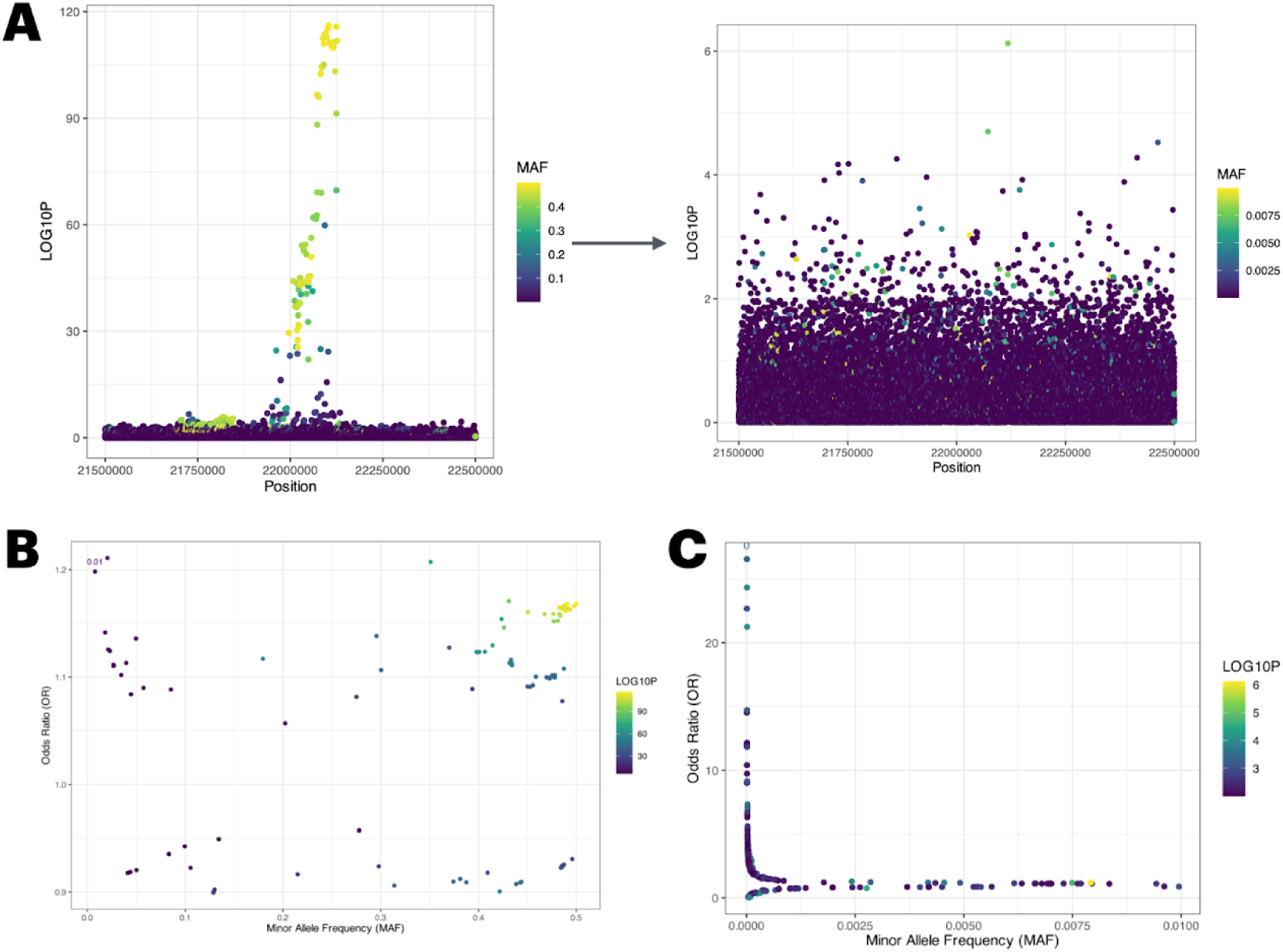
CAD association at locus 9p21.3 utilizing whole genome sequencing in UK Biobank. Coronary artery disease (CAD) association testing was performed using Regenie in individuals of European ancestry from the UK Biobank whole-genome sequencing cohort. SNPs with minor allele count <20 were excluded. The final analysis included 54,438 cases and 403,516 controls. (A) Locus zoom plot of the 9p21.3 region, with genomic position on the x-axis and −log₁₀(p-value) on the y-axis; points are colored by minor allele frequency (MAF). The plot on the right zooms in on variants with MAF <1%. (B) Scatterplot of MAF versus odds ratio for SNPs with p-values <1×10⁻⁶. (C) Scatterplot of MAF versus odds ratio for SNPs with p-values <1×10⁻² and MAF<1%.

**Supplementary Figure 2:**
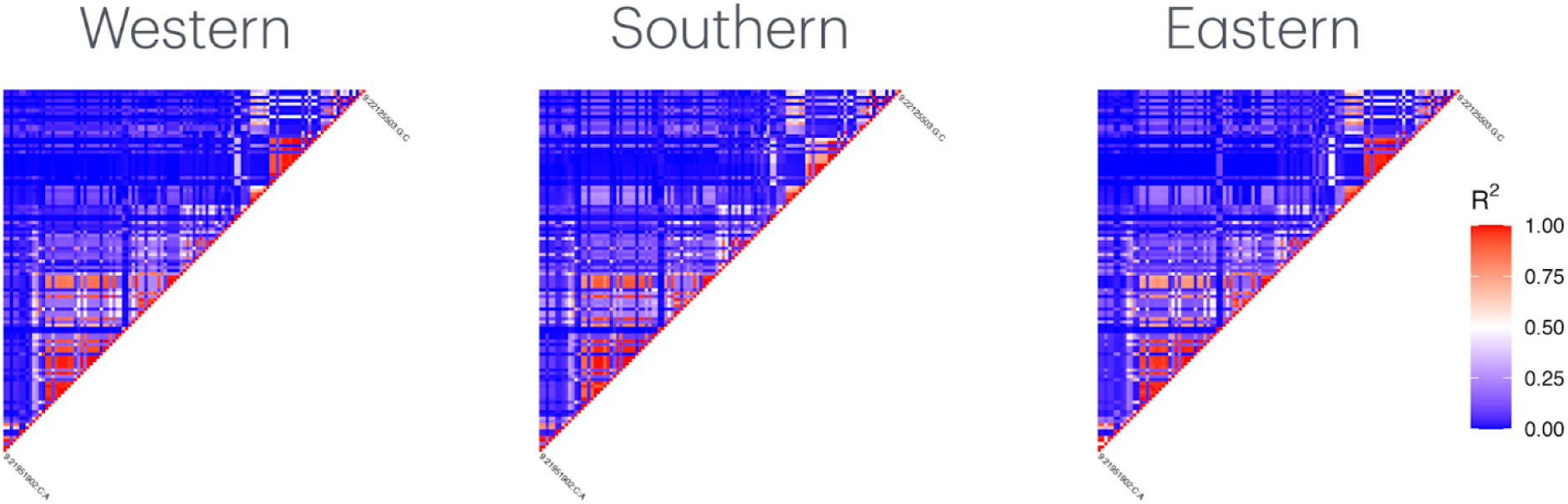
Linkage disequilibrium heat maps of CAD significant SNPs in AWI-Gen. Linkage disequilibrium (LD) reflects in-sample r2 in Western, Southern, and Eastern regions in AWI-Gen cohort. SNPs shown in the LD heatmaps reached genome-wide significance in at least one studied ancestry group and has a minor allele frequency >1% in all groups.

**Supplementary Figure 3:**
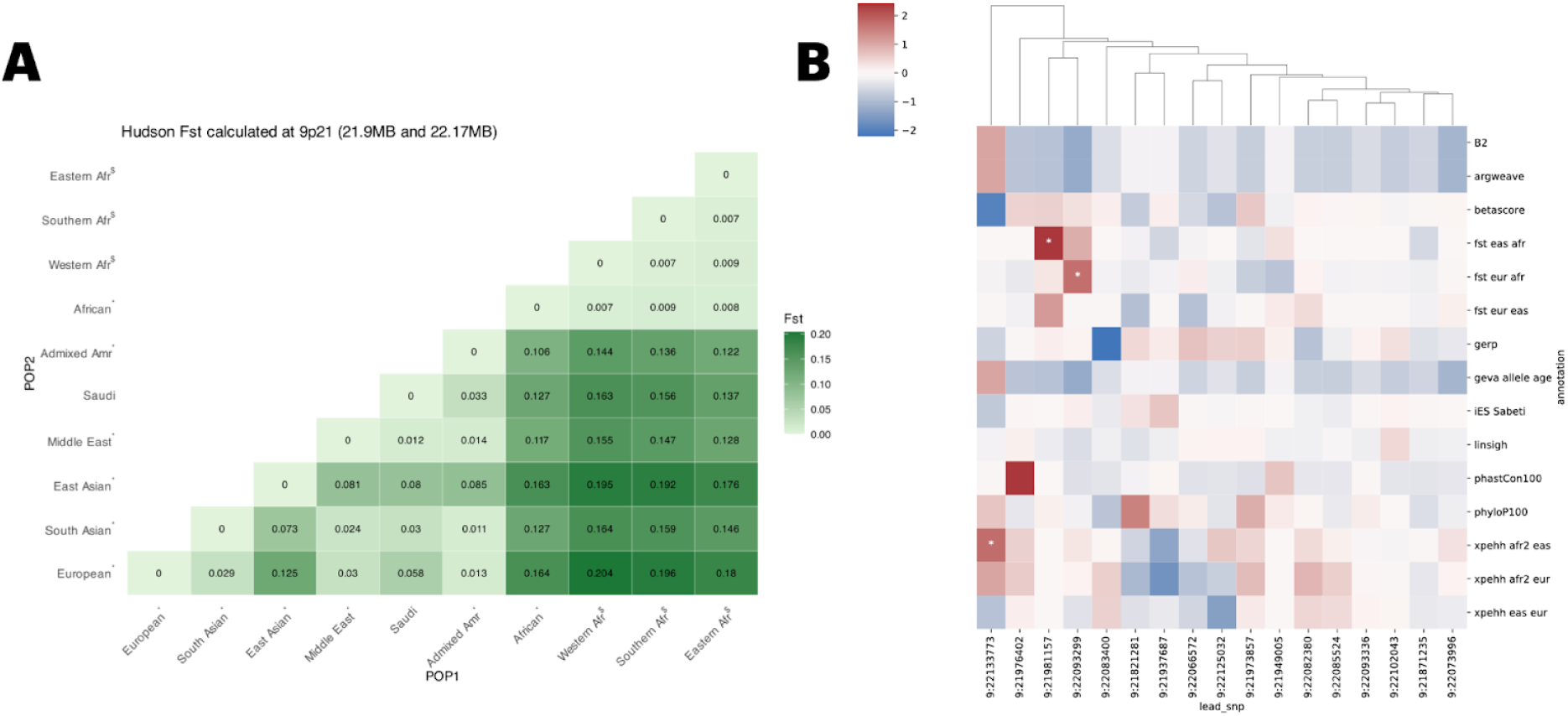
Population differentiation and evolutionary enrichment at locus 9p21.3. (**A**) Pairwise Hudson’s Fst values between ancestry groups at the primary association hotspot of 9p21.3 (21.9–22.17 Mb). Asterisks (*) denote ancestry groups from the UK Biobank, dollar signs ($) indicate groups from the AWI-Gen cohort, and “Saudi” refers to the King Faisal Specialty Hospital and Research Centre cohort. (**B**) Heatmap showing enrichment of evolutionary signatures—including metrics related to selection, conservation, and constraint—among independent SNPs at 9p21.3, as estimated using the GSEL package. Homozygous European summary statistics were used as input into GSEL. Asterisks indicate SNPs with statistically significant enrichment, as defined in Methods.

**Supplementary Figure 4:**
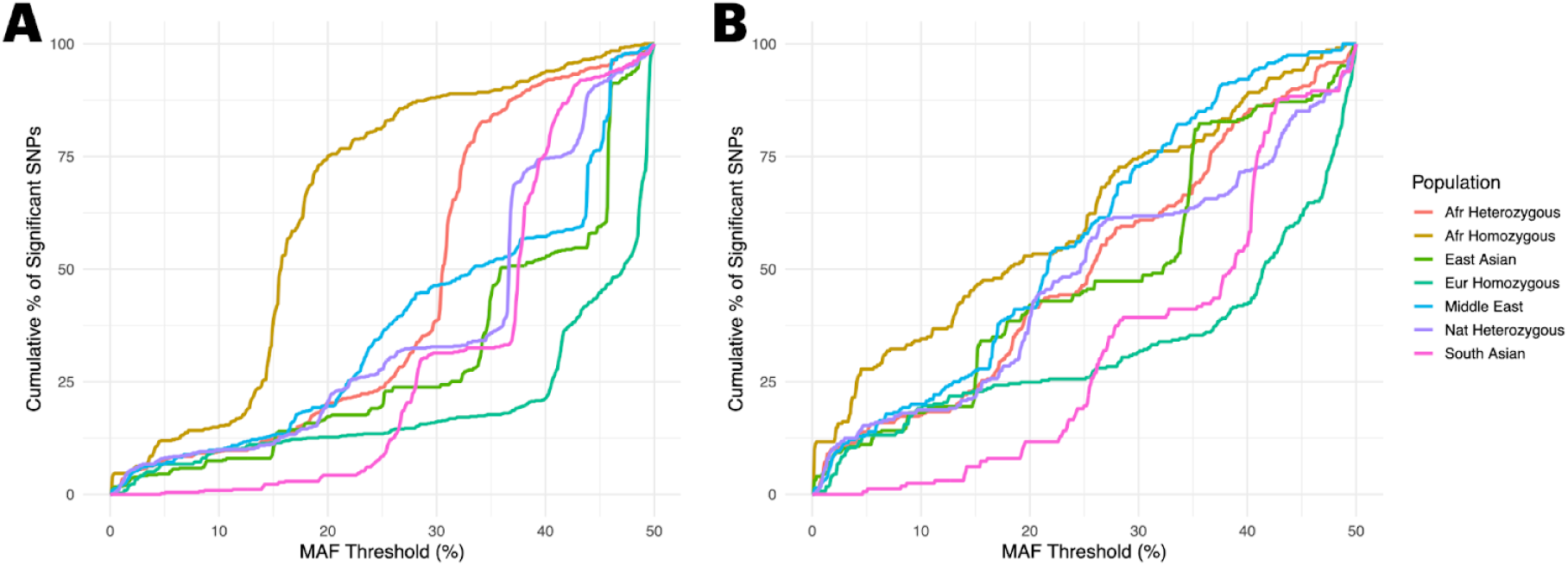
Cumulative Distribution of MAF for CAD-Associated SNPs Across Populations. Cumulative distribution plots showing the percentage of CAD-associated SNPs with MAF less than or equal to a given threshold, stratified by population. (A) Distribution for all 667 genome-wide significant SNPs across the full 9p21.3 region. (B) Distribution for the 298 significant SNPs located within the primary hotspot (21.9–22.13 MB).

**Supplementary Figure 5:**
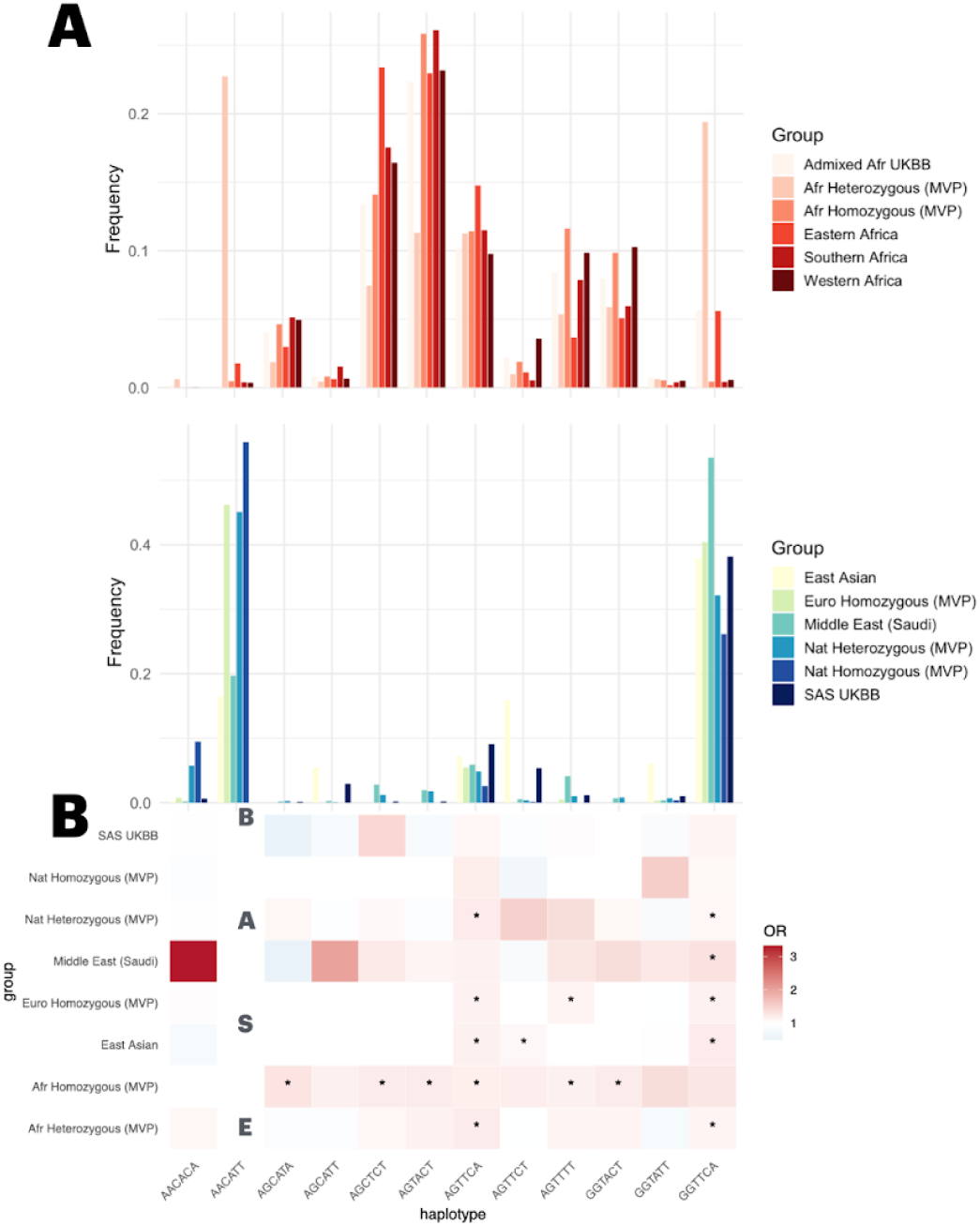
Replication of haplotype analysis from Tcheandjieu, et al. (2022) across ancestry groups. Haplotype analysis was performed across multiple cohorts—including MVP, BBJ, King Faisal Specialist Hospital and Research Centre (KFSHRC), AWI-Gen, and UK Biobank (UKBB)—capturing individuals of African, European, East Asian, Middle Eastern, and South Asian ancestry. Haplotype frequencies and associations with CAD were estimated using the haplo.stats package in R. The six SNPs used in this analysis were previously described in Tcheandjieu et al. (2022). Haplotypes with very low frequency were removed for clarity. (**A**) Top panel shows haplotype frequencies in African-relevant groups. Eastern, Southern, and Western African samples are from AWI-Gen, while “heterozygous” and “African homozygous” refer to local ancestry (LAI) groups in MVP. The bottom panel shows frequencies in BBJ, Middle Eastern individuals (KFSHRC), European and Indigenous American LAI groups (MVP), and South Asians (UKBB). (**B**) Haplotype associations with CAD for cohorts where phenotype data were available. Association models included sex and the first five principal components as covariates. AACATT was used as the reference haplotype; however, it has very low frequency in the African homozygous group, resulting in unstable regression estimates. Squares are colored by odds ratio, and stars denote Bonferroni-corrected p-values < 0.05.

**Supplementary Figure 6:**
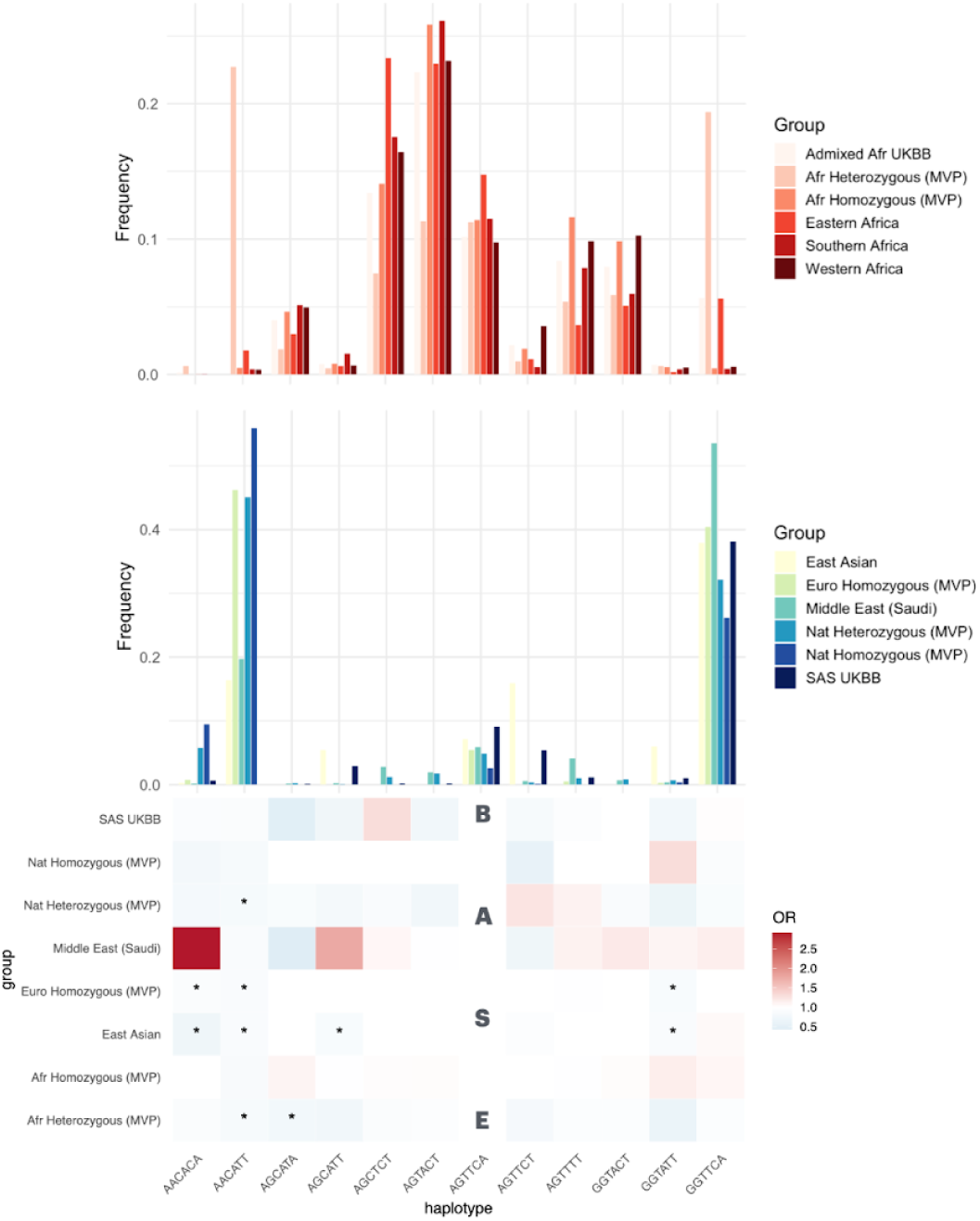
Replication of haplotype analysis from Tcheandjieu, et al. (2022) across ancestry groups. This figure mirrors the structure and cohorts described in Figure S2.3 (see legend for full details), with the only difference being the reference haplotype used in the association models. Here, AGTTCA was used as the baseline haplotype instead of AACATT. AGTTCA was chosen due to its relatively consistent frequency across ancestry groups, allowing for more stable regression estimates.

**Supplementary Figure 7:**
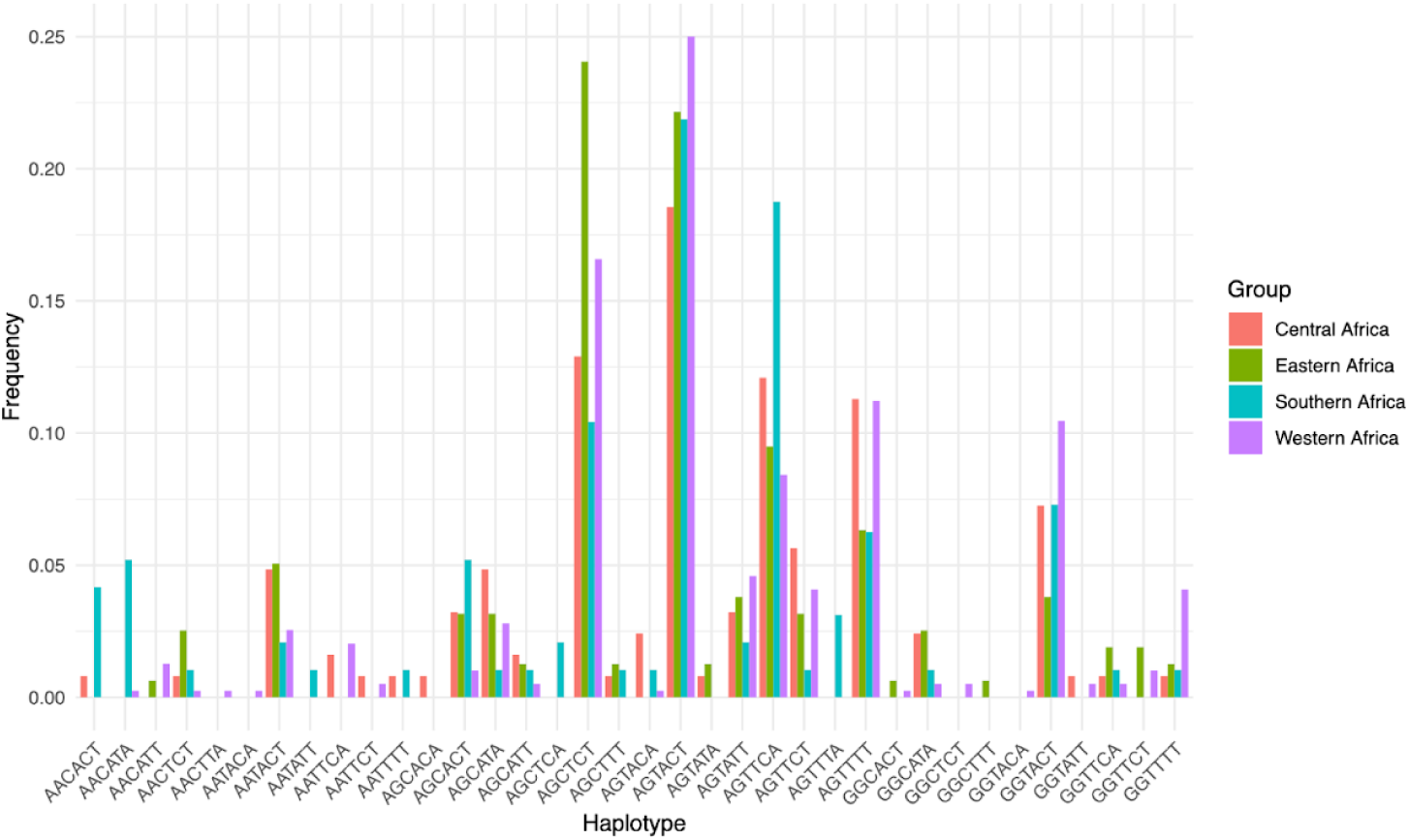
6 SNP haplotype frequencies in H3Africa whole genome sequencing. This figure reflects the same 6 SNP used in Fig S3 and S4. Haplotypes were directly observed using the phased data, and therefore were not estimated.

**Supplementary Figure 8:**
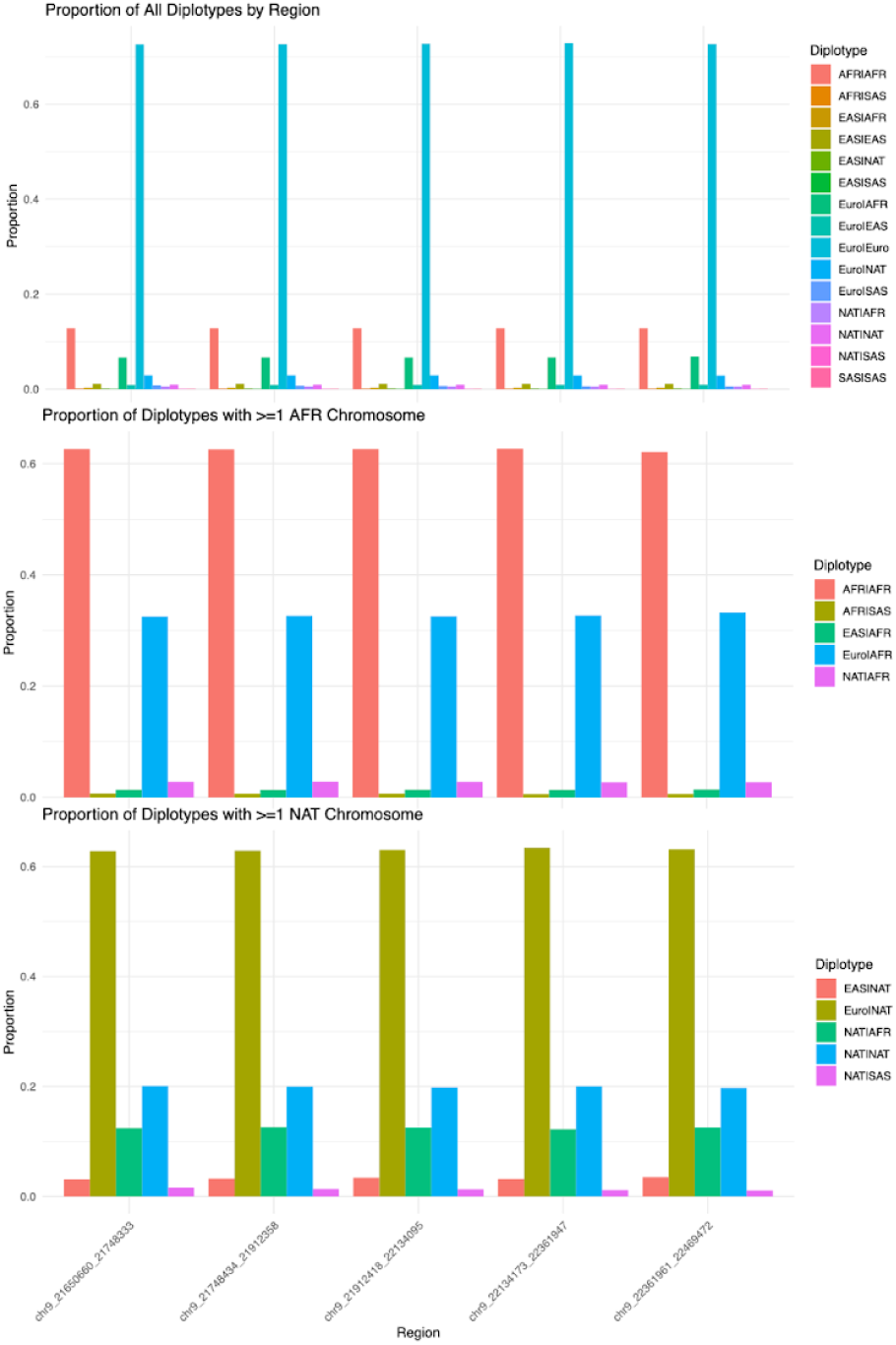
Local ancestry diplotype frequencies at locus 9p21.3. Local ancestry inference was performed across the entire MVP cohort, as described in Methods. Ancestries included in the inference were African (Afr), Indigenous American (Nat), European (Euro), East Asian (Eas), and South Asian (Sas). The 9p21.3 region was divided into five genomic intervals to capture local variation across the locus. The top panel shows diplotype frequencies at each of the five intervals. The middle panel shows diplotype frequencies among individuals carrying at least one African-ancestry chromosome. The bottom panel shows diplotype frequencies among individuals carrying at least one Indigenous American-ancestry chromosome.

**Supplementary Figure 9:**
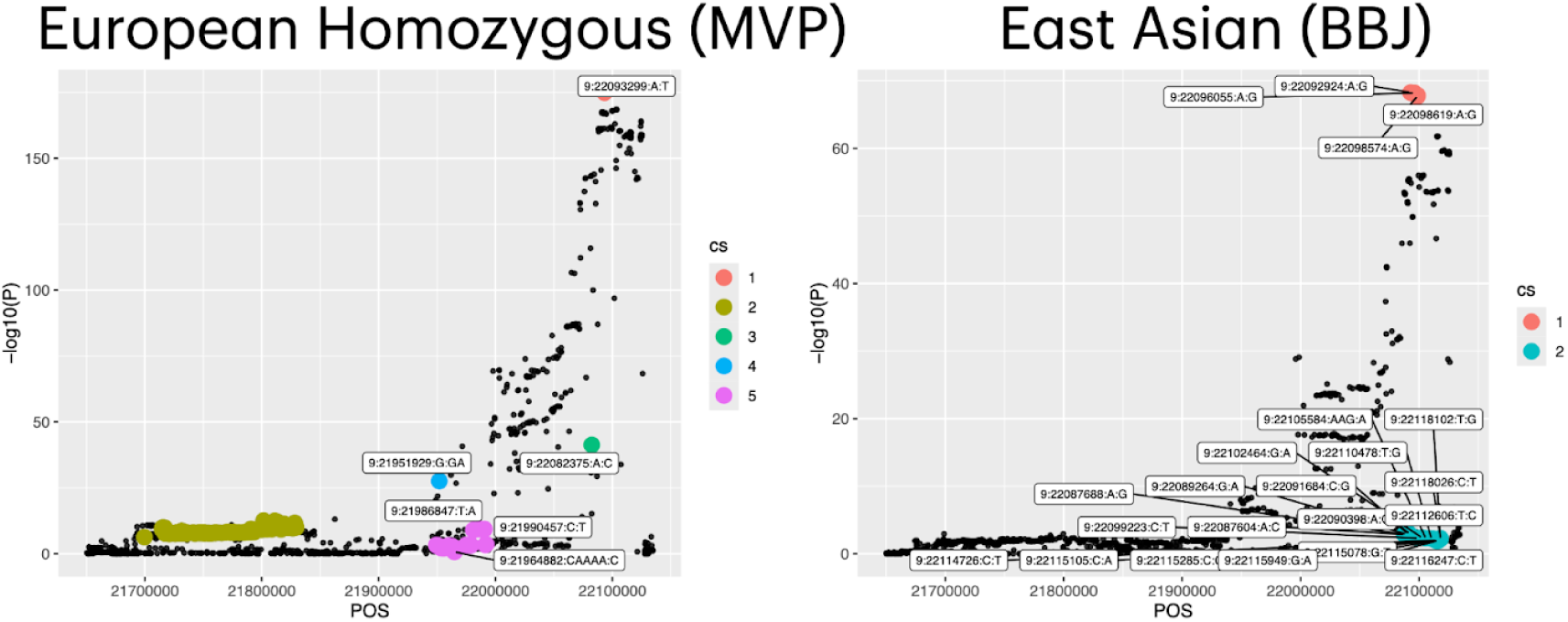
**Fine-mapping of locus 9p21.3 in European and East Asian ancestries using SuSiE-R**. Fine-mapping was performed using SuSiE-R with summary statistics and in-sample LD to identify credible sets of putative causal variants at the 9p21.3 locus. Only common variants (MAF > 5%) were included, consistent with the filtering used in the conditional analyses. The left panel shows results from the European homozygous group defined by local ancestry inference (LAI) in MVP, and the right panel shows results from East Asian individuals in BioBank Japan (BBJ). Variants included in each credible set are color-coded, representing distinct credible sets inferred by the SuSiE model.

**Supplementary Figure 10:**
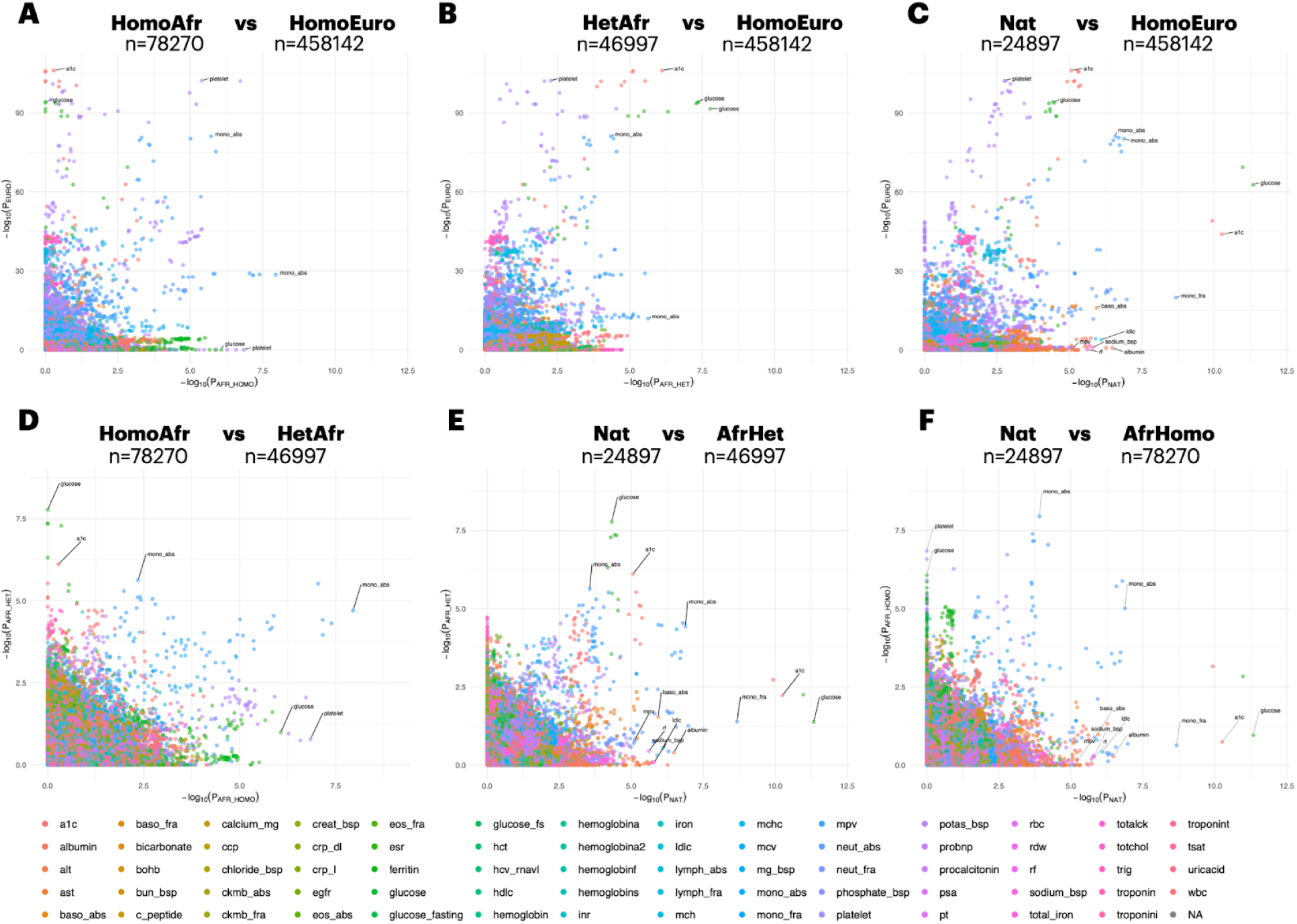
Comparison of phenome-wide association on various available blood markers across local ancestry groups in MVP. Phenome-wide association was conducted on available blood markers in MVP, stratified by local ancestry at 9p21.3, focusing on homozygous and heterozygous ancestry groups for African, European, and Indigenous American (Nat) chromosomes. Nat group includes both heterozygous and homozygous Nat chromosomes in order to increase sample size. Panels (**A-F**) shows a pairwise comparison of −log₁₀(p-values) between two groups. Variants with minor allele frequency <5% were excluded. Phenotypes with fewer than 500 samples were also removed. Points are colored by phenotype category (blood marker), and selected points are labeled if they reached genome-wide significance in the European group or nominal significance in other groups. To reduce clutter, only the top SNP for each phecode was selected for labeling. Full association details are provided in the Methods section.

**Supplementary Figure 11:**
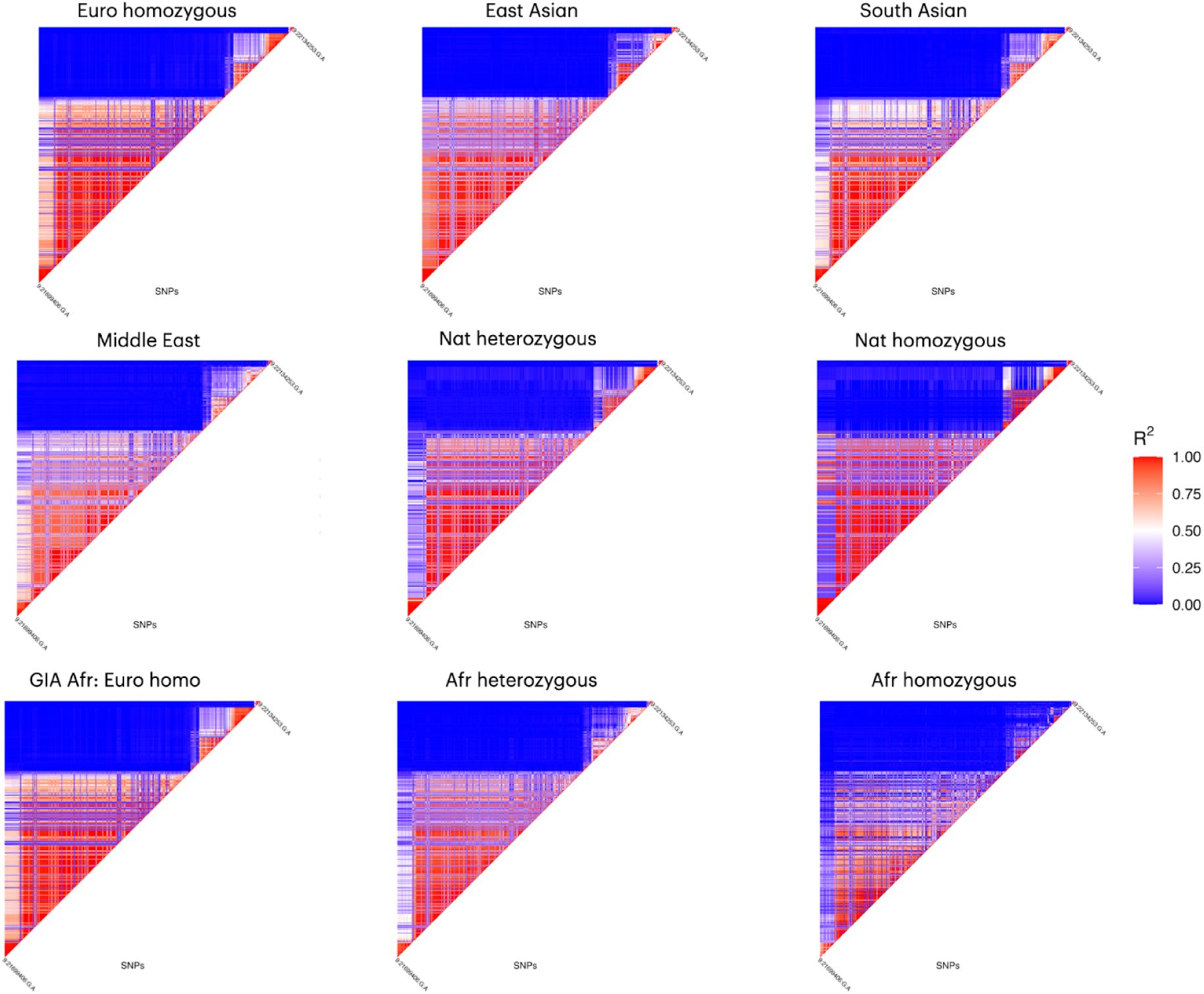
Linkage disequilibrium heatmaps of PheWAS-significant SNPs across ancestries. Linkage disequilibrium (LD) heatmaps show in-sample r² across ancestry groups. SNPs included in the analysis reached genome-wide significance in the European homozygous group, had a minor allele frequency (MAF) ≥1% in all ancestries, and were required to have LD information available in every group. A total of 482 SNPs met these criteria.

**Supplementary Figure 12:**
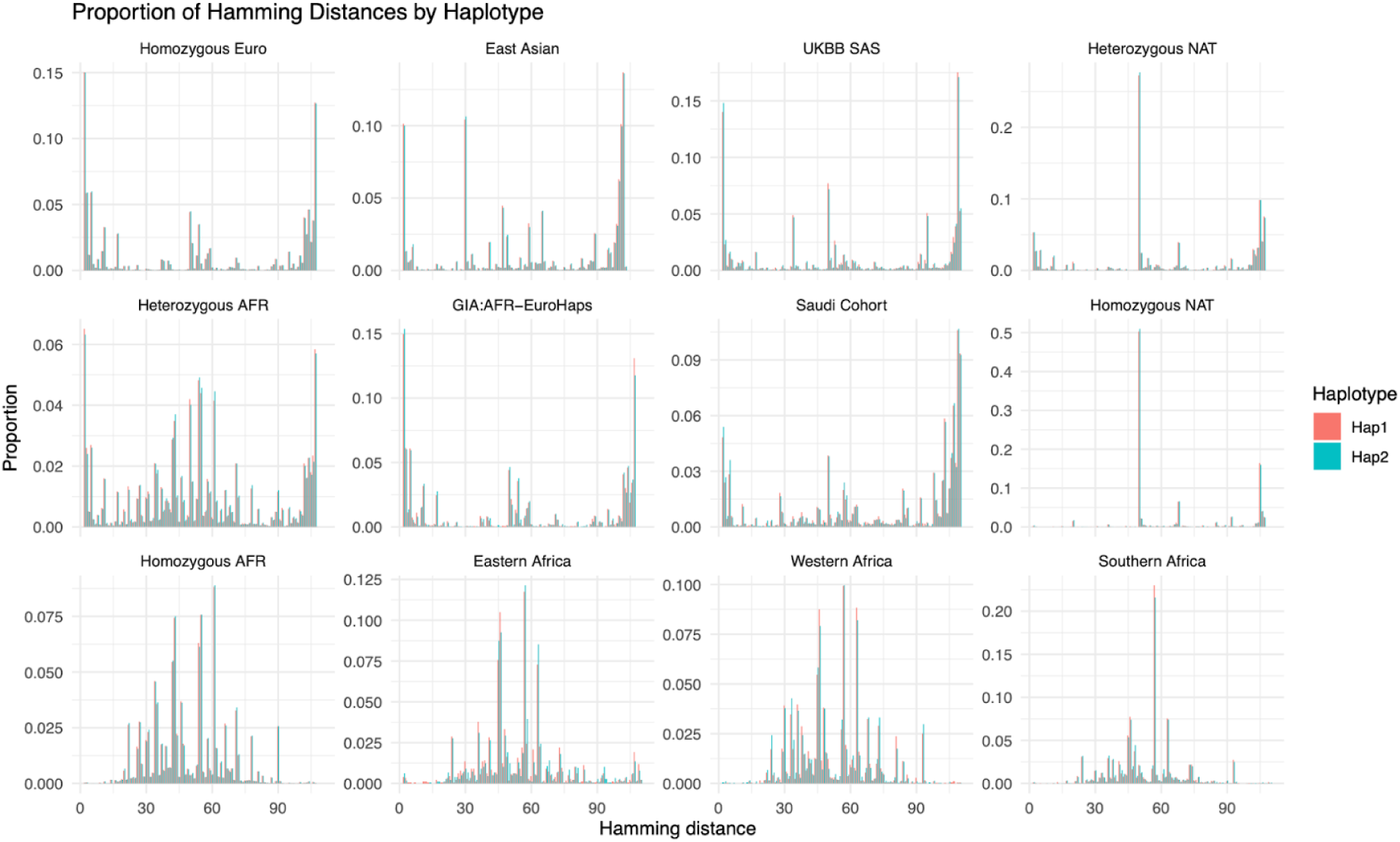
Hamming distance across cohorts at locus 9p21.3. Hamming distances were calculated for each individual haplotype (hap1 and hap2) relative to the theoretical non-risk haplotype. Panels show the distribution of hap1 and hap2 distances within each population group, with the two haplotypes distinguished by color.

